# A Pleiotropic Map of Brain Imaging Genetics Reveals Biologically Distinct Latent Endophenotypes

**DOI:** 10.64898/2026.04.27.26351743

**Authors:** Upasana Bhattacharyya, Jibin John, Zhong Yuanxin, Michael Preuss, Tian Ge, Todd Lencz, Max Lam

## Abstract

Genomewide association studies (GWAS) of brain scans are complicated by the large number and high collinearity of the available image-derived phenotypes (IDPs). Here, we present DIMPLE-GWAS (Dimensionality reduction and Integrated Multi-Phenotype Landscape Explorer for GWAS), a dimensionality-reduction framework designed to identify latent genetic architecture across high-dimensional pleiotropic phenotypes. This approach, applied to ∼4000 IDPs from ∼33K European ancestry participants in the UK Biobank, yielded 25 biologically interpretable latent phenotypes; this structure was validated in the independent ABCD cohort. The DIMPLE-GWAS clusters demonstrated substantially greater heritability than the input IDPs and yielded greater power for locus discovery, including 104 genomewide-significant loci not reported in prior GWAS of individual IDPs. These genetically defined phenotypes only partially aligned with conventional brain atlas boundaries based on gyral, cytoarchitectonic, or functional features. Instead, they revealed distinct patterns of brain organization and novel genetic relationships with neurologic, psychiatric, cognitive, and behavioral phenotypes.

## INTRODUCTION

Neuroimaging genetics has long held the promise of revealing novel biological insights into the biology of the brain and its relationship to cognitive, behavioral, psychiatric, and neurological phenotypes^1^. After initial progress was made through large-scale collaborative efforts focusing on specific brain regions^2^, the UK Biobank (UKBB) provided the first release of comprehensive, large-scale neuroimaging data^3,4^. However, the very richness of this dataset presents a formidable analytical challenge. The high dimensionality of the UKBB neuroimaging datasets, comprising thousands of imaging-derived phenotypes (IDPs), creates a massive multiple testing burden that can obscure true biological signals. Standard univariate GWAS, which analyzes each IDP independently, fails to capitalize on the correlated genetic effects that are the hallmark of pleiotropic brain IDPs^5^. At the same time, classic anatomically-derived brain atlases do not necessarily respect the underlying genetics of regional brain variation^6^. While multivariate approaches have been developed to reduce dimensionality and identify shared signals across brain IDPs^7^, these efforts have not yet clearly resolved the latent endophenotypes underlying the shared genetic signals. Additionally, prior studies have not yet applied multitrait approaches comprehensively across all IDPs, which include modalities ranging from structural to diffusion to functional neuroimaging.

The present study introduces and applies a novel multitrait analytic framework, designed to identify genetically-derived latent constructs from high-dimensional phenotypes such as neuroimaging IDPs: DIMPLE-GWAS (Dimensionality reduction and Integrated Multi-Phenotype Landscape Explorer for GWAS). DIMPLE-GWAS is an unsupervised, data-driven workflow that employs Uniform Manifold Approximation and Projection (UMAP)^8^, a powerful non-linear dimensionality reduction technique, to embed thousands of IDPs into a lower-dimensional space based on the similarity of their genome-wide SNP association profiles. Subsequently, Hierarchical Density-Based Spatial Clustering of Applications with Noise (HDBSCAN)^9^ identifies stable, dense clusters within this embedding, grouping IDPs that share a common genetic architecture. The central hypothesis of this work is that these genetically-defined clusters represent more robust and biologically coherent phenotypes than their individual constituent IDPs.

By applying DIMPLE-GWAS to GWAS summary statistics from thousands of IDPs from the UK Biobank BIG-40 database^4^ (https://open.oxcin.ox.ac.uk/ukbiobank/big40/), this study aims to: (1) create a comprehensive, data-driven pleiotropic map of the human brain’s genetic architecture; (2) demonstrate that this clustering approach enhances statistical power for the discovery of novel genetic loci; (3) functionally annotate the identified loci and clusters to uncover the specific biological pathways, cell types, and developmental stages through which genetic variation shapes the brain; and (4) leverage these genetically-defined brain clusters as instruments to investigate shared etiology and causal relationships with a wide array of psychiatric, neurological, and behavioral traits using bivariate linkage disequilibrium score regression (LDSC) and Mendelian randomization (MR). This work provides a powerful new lens through which to view the genetic organization of the brain, moving beyond single-trait associations to a systems-level understanding of how shared genetic mechanisms contribute to both normal variation and disease.

## RESULTS

### DIMPLE-GWAS clustering

#### Primary clustering of the full IDP set

As summarized in **Figure 1**, the first step of our workflow was to download UK Biobank IDPs across structural (including thickness, area, volume, and intensity measures), diffusion, and functional MRI modalities, and then filter out IDPs with heritability that did not significantly differ from zero (as determined by LDSC). We then subjected the remaining 2,315 IDPs with non-zero heritability to the DIMPLE-GWAS pipeline (employing UMAP for dimensionality reduction and HDBSCAN for clustering; see Methods for details). The optimal final configuration (as determined by an automated optimisation of hyperparameters after a brute-force grid search, see Methods for details) yielded 15 clusters (excluding 153 (6.7%) IDPs labelled as noise). This solution achieved mean silhouette = 0.612 (considered “reasonable”)^10^ with a corresponding fit score = 0.846. Each cluster contained between 42 and 459 IDPs (median=121).

**Figure 1.**
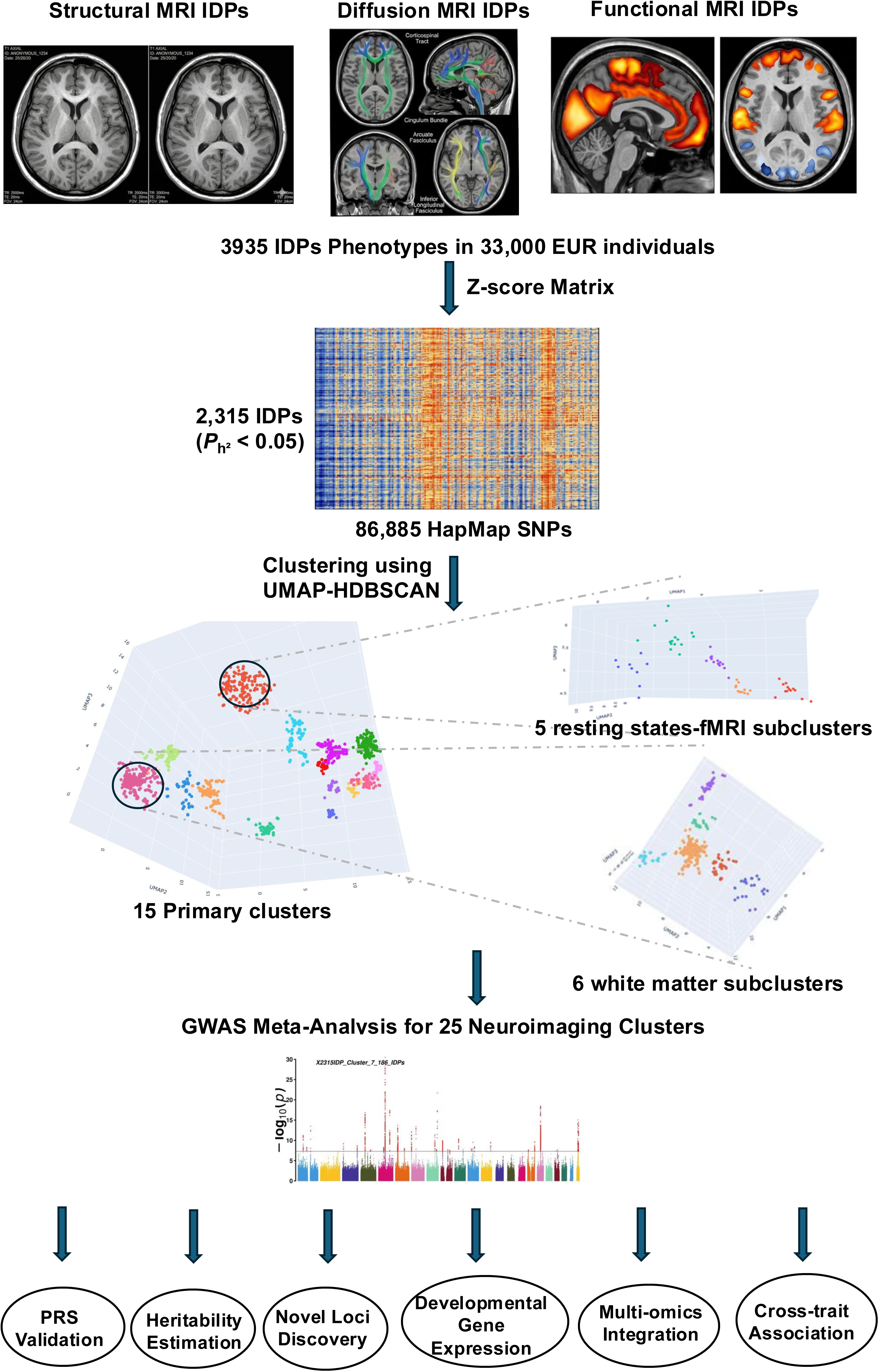
Workflow for constructing a pleiotropic map of brain imaging phenotypes. Overview of the DIMPLE-GWAS analytical pipeline. Starting from 3,935 imaging-derived phenotypes (IDPs) measured across 33,000 European-ancestry individuals from the UK Biobank, Z-score matrices were computed and 2,315 IDPs with significant heritability (PL² < 0.05) were retained. Genetic similarity among IDPs was quantified using 86,885 HapMap SNPs. Dimensionality reduction and unsupervised clustering via UMAP-HDBSCAN identified 15 primary clusters spanning structural MRI, diffusion MRI, and functional MRI IDPs. Two-stage secondary clustering further resolved 6 white matter subclusters and 5 resting-state fMRI subclusters, yielding 25 genetically-defined neuroimaging clusters in total. These 25 clusters were taken forward to GWAS meta-analysis, followed six downstream analyses: polygenic risk score (PRS) validation, heritability estimation, novel loci discovery, developmental gene expression profiling, multi-omics integration, and cross-trait genetic association.

As shown in **Figure 2A** (and detailed in **Supplementary Table 1a**), the resulting solution yields readily interpretable, biologically coherent clusters defined both by imaging modality and regional specificity. Most notably, volumetric and surface area (SA) measures of cortical structures formed shared clusters delineated primarily by lobe; separate clusters contained volume/SA IDPs specific to occipital, temporal, parietal, and (orbital) frontal lobes, with one cluster specific to the volume/SA of the cingulate cortex. Unexpectedly, two clusters were identified that extended across lobar boundaries: one which captured volume/SA of the posterior ventral surface of the brain, ranging from the lingual gyrus of the occipital cortex along the fusiform gyrus to the entorhinal and parahippocampal cortex of the temporal lobe; the other contained IDPs representing volume/SA measures of dorsal structures ranging from frontal pole and dorsal prefrontal cortex to the superior occipital cortex (and including somatomotor cortex, supramarginal cortex, superior temporal gyrus, and angular gyrus). Notably, IDPs representing total cortical volume also were included in this dorsal volume/SA cluster.

**Figure 2.**
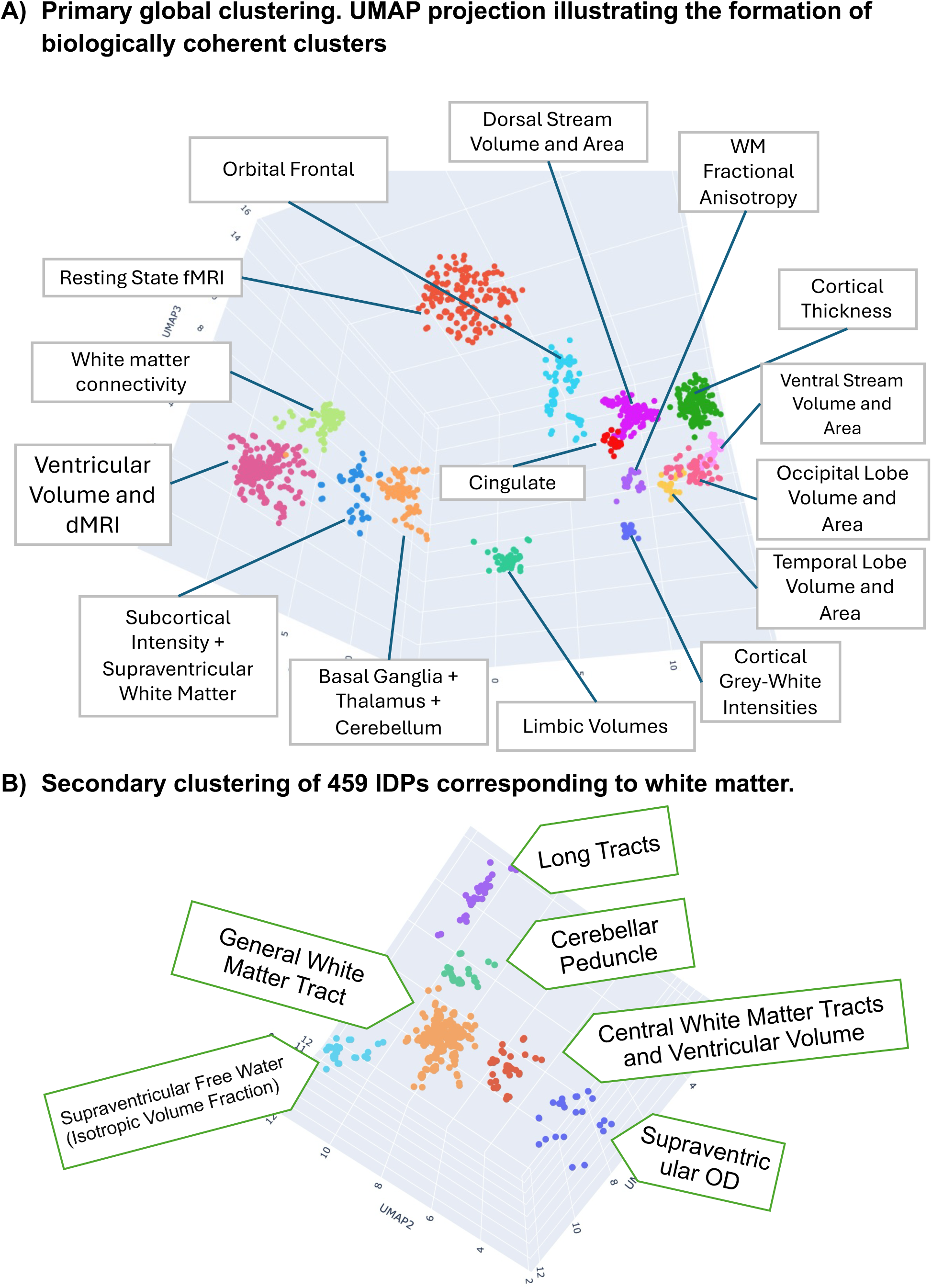
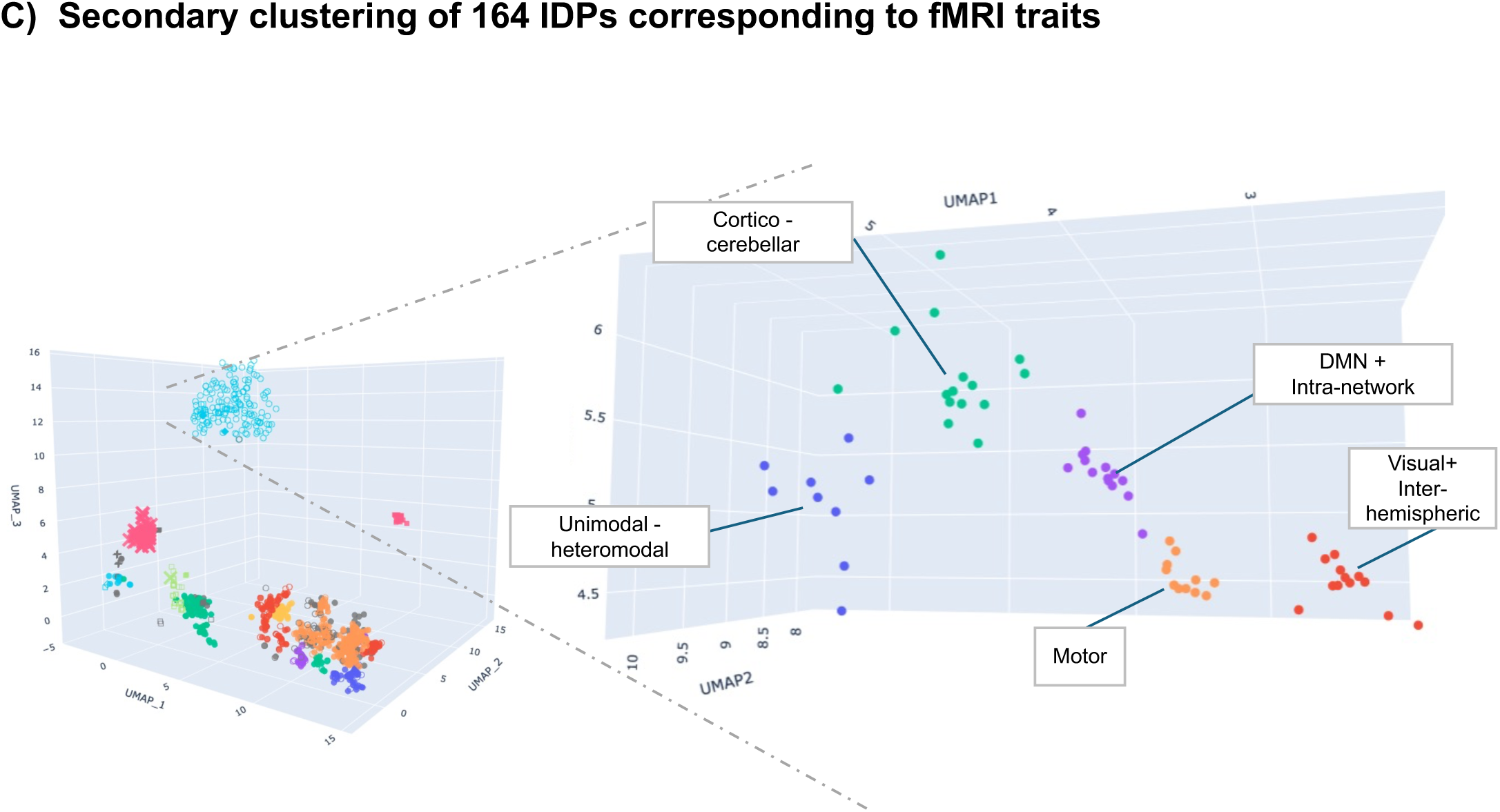
Primary and secondary multidimensional clustering of neuroimaging-derived phenotypes. **A.** Primary global clustering. UMAP projection illustrating the formation of 15 biologically coherent clusters from 2,315 heritable IDPs. Each point represents a single IDP, coloured by cluster assignment. The spatial separation of clusters reflects distinct patterns of genetic covariation across structural MRI, diffusion MRI, and resting-state functional MRI phenotypes. **B.** Secondary clustering of 459 IDPs corresponding to white matter diffusion phenotypes. UMAP re-embedding of white matter IDPs resolved 6 subclusters, capturing finer-grained genetic architectures within the white matter domain. Points are coloured by subcluster identity; UMAP coordinates reflect the secondary embedding space. **C.** Secondary clustering of 164 IDPs corresponding to resting-state fMRI traits. Re-embedding of functional connectivity IDPs resolved 5 subclusters, delineating genetically distinct functional network components. Points are coloured by subcluster identity.

While the volume/SA IDPs were regionally divided as described above, most cortical thickness IDPs formed a single cluster, encompassing all cortical regions. Similarly, cortical gray/white intensity measures formed a single cluster, as did all functional neuroimaging measures (primarily resting-state fMRI, as many task-based fMRI measures either did not cluster or were eliminated from the analysis due to non-significant heritability). Subcortical regional volumes were divided into three clusters, relatively distant in UMAP space from the cortical clusters: 1) limbic (hippocampus/amygdala) volumes; 2) ventricular volumes; 3) volumes of all other deep structures (basal ganglia/thalamus/cerebellum/brain stem). IDPs derived from diffusion MRI (dMRI) were also divided across three clusters: 1) intracellular volume (ICVF) and fractional anisotropy (FA) measures from all regions (except the fornix) fell into a single cluster (labeled white matter connectivity, WMC) that contained no other IDPs; 2) measures of orientation dispersion (OD) and mode of anisotropy (MO) from several fiber tracts, primarily located in the supraventricular region, formed a cluster along with IDPs capturing the signal intensity of subcortical structures; and 3) a widely distributed set of dMRI-derived IDPs (including MO and OD from most tracts, as well as all regional measures of free water and diffusion eigenvalues L1, L2, and L3). The latter formed a very large cluster (459 IDPs) along with the ventricular volumes noted above.

#### Secondary clustering within diffusion MRI and functional MRI feature clusters

To assess finer-grained structure within the two clusters containing 459 widespread dMRI measures and 164 widespread fMRI IDP measures, respectively, we performed secondary clustering using the UMAP/HDBSCAN approach on these IDPs only. The dMRI subset exhibited strong internal structure, achieving a “reasonable” silhouette score = 0.526 and a fit score = 0.956. Only 16/459 IDPs were labelled as noise (3.4%), and the remaining 443 IDPs were assigned to 6 clusters as follows (**Figure 2B; Supplementary Table 1b)**: 1) Supraventricular OD; 2) Supraventricular Free Water; 3) Central White Matter Tracts and Ventricular Volumes; 4) Cerebellar Peduncle (including 4th ventricular volume); 5) White Matter Long Tracts (e.g., medial lemniscus and corticospinal tract); and 6) a large cluster of regionally distributed, diverse white matter IDP measures (referred to as “General White Matter Tract”). In downstream analyses, these six secondary clusters were utilized rather than the large cluster of all 459 IDPs.

On the other hand, for the large fMRI cluster, the optimal configuration labelled a majority of observations as noise (102/164; 62.19%), leaving 62 IDPs assigned to 5 clusters with small cluster sizes (9–15, median 13). Despite a reasonable mean silhouette score of 0.622, the fit score (0.338) indicated that separable structure within this subset is comparatively weak and the derived subclusters should be interpreted as exploratory. Consequently, in downstream analyses, we examined both the full cluster of 164 IDPs as well as the 5 subclusters, which were characterized as follows (**Figure 2C; Supplementary Table 1c**): 1) resting state fMRI (rs-fMRI) IDPs linking unimodal and heteromodal cortex; 2) rs-fMRI IDPs representing visual cortex and inter-hemispheric networks; 3) cortico-cerebellar rs-fMRI networks; 4) default mode network and other intra-network rs-fMRI connectivity; and 5) rs-fMRI motor networks. Thus, the total number of clusters that will be subjected to downstream analyses is 25: 5 rs-fMRI subclusters, 6 dMRI subclusters, and 14 of the original 15 primary clusters (i.e., excluding the large dMRI cluster from the primary analysis). These clusters, with abbreviations that will be used throughout this text, are listed in **Supplementary Table 1d**. As described in the Methods, cluster-level summary statistics were computed for each of these 25 clusters using genomic PCA^11^, and replicability was tested using cluster-derived polygenic scores computed in the ABCD dataset.

### Heritability analysis

Cluster-level heritability estimates derived from LDSC were consistently higher than the heritability of constituent individual IDPs across all identified DIMPLE-GWAS clusters, indicating enhanced capture of genetic signal through dimensionality reduction. For every cluster examined, the cluster heritability (h^2^) exceeded the mean and median heritability of the individual IDPs assigned to that cluster, often by a substantial margin (**Figure 3**). The median difference between cluster-level heritability and mean within-cluster heritability was 0.075.

**Figure 3.**
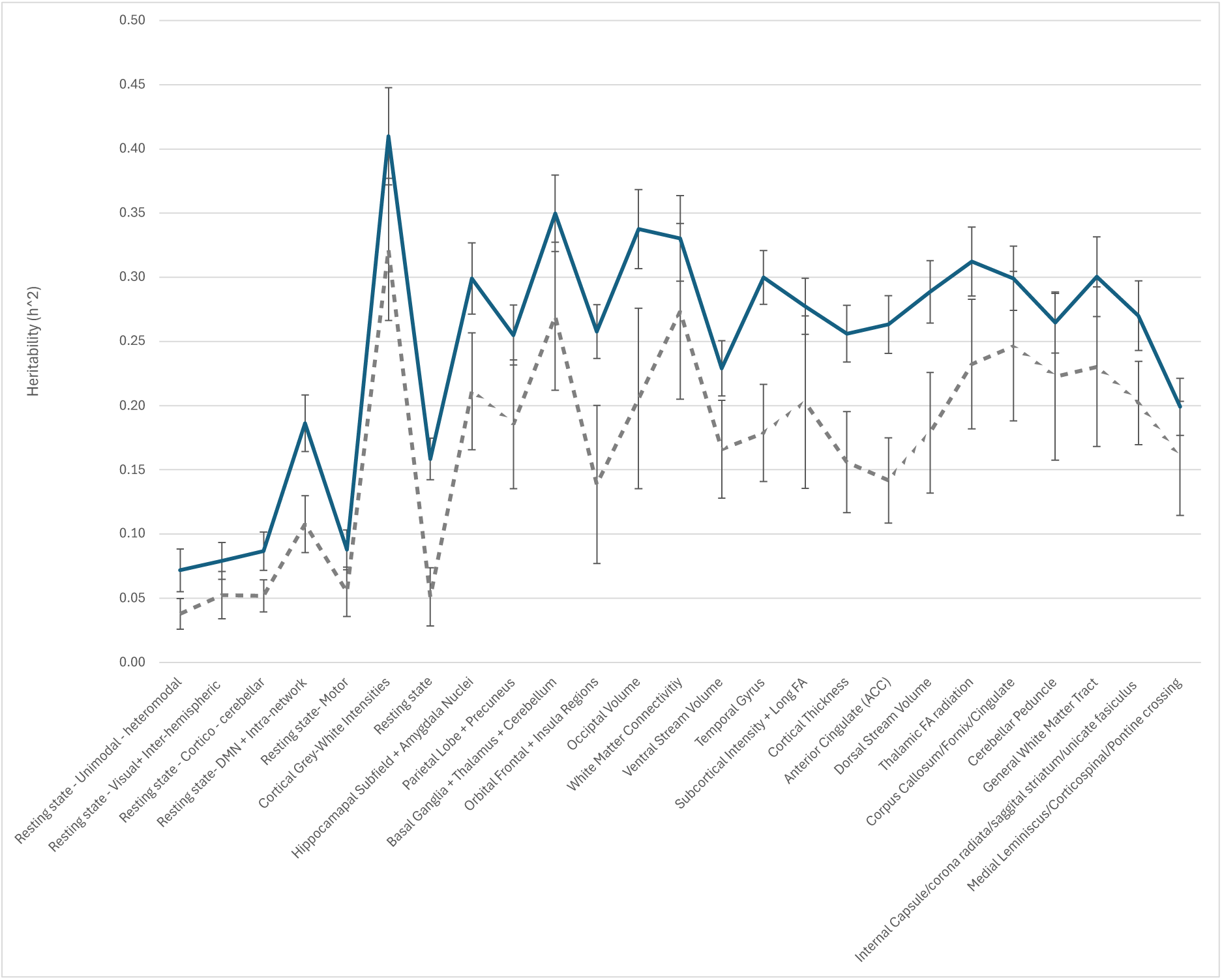
LDSC heritability estimates for DIMPLE-GWAS clusters compared with constituent individual IDPs. Linkage disequilibrium score regression (LDSC) SNP-heritability estimates (h²) for each of the 25 DIMPLE-GWAS clusters (shown in blue colour) plotted alongside the distribution of h² values for their constituent individual IDPs (shown in grey). Cluster-level h² estimates are consistently higher than the median IDP-level estimates within the same cluster, demonstrating that aggregation of genetically coherent IDPs into clusters amplifies detectable heritability. Error bars represent standard errors.

### Validation of DIMPLE-GWAS clusters using ABCD polygenic scores

To evaluate the validity and generalizability of the neuroimaging clusters derived from DIMPLE-GWAS performed on UK Biobank data, we performed an external validation analysis using polygenic risk scores (PRS) calculated in the Adolescent Brain Cognitive Development (ABCD) cohort^12^. Specifically, we assessed whether the PRS derived from summary statistics of each cluster could predict phenotypic variance of the corresponding imaging-derived phenotypes^13,14^ within the independent ABCD dataset. Of the >51,000 IDPs examined in ABCD dataset, the IDP with the strongest correlation to the PRS for each of the primary DIMPLE clusters (except for the main resting state fMRI cluster) was a phenotype corresponding to the DIMPLE cluster content, with p-values ranging from 2.56×10^-11^ to 1.36×10^-81^. For example, the PRS derived from the temporal lobe volume/SA DIMPLE cluster (TMP) was most strongly associated with ABCD variable “smri_area_cdk_mdtmlh” (Cortical area in mm^2^ of APARC ROI lh-middletemporal; R^2^=0.22; p=1.13×10^-24^). We compiled a table listing all correlation values between the PRS for each DIMPLE cluster and corresponding ABCD IDPs (**Supplementary Table** 2). The p-values for these correlations were transformed to −log₁₀(p) scores and displayed in **Figure 4**, which demonstrates that each of the primary clusters and white matter secondary clusters demonstrated strong associations with corresponding ABCD phenotypes, providing fully independent support for the DIMPLE-GWAS approach.

**Figure 4.**
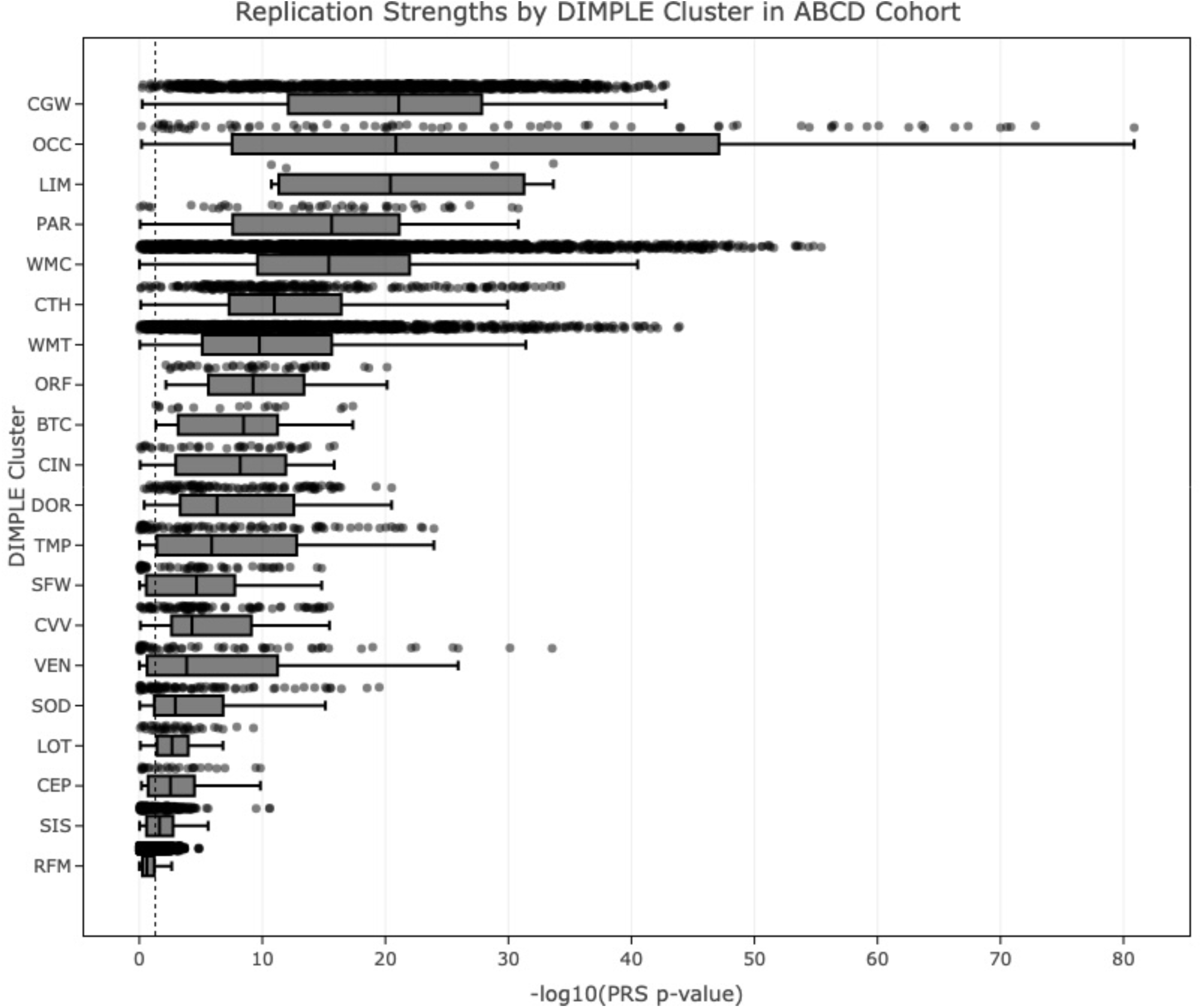
Polygenic risk score (PRS) validation of DIMPLE-GWAS clusters in the independent ABCD Study. Association strengths (expressed as −log₁₀ P-values) between UK Biobank-derived cluster PRSs and their top correlated imaging phenotypes measured in the Adolescent Brain Cognitive Development (ABCD) cohort (NL=L4,264 adolescents).

Due to differences in phenotype construction between ABCD and the UK Biobank for resting state fMRI IDPs, it was difficult to specify the appropriate corresponding ABCD IDPs for the secondary DIMPLE-GWAS fMRI clusters; therefore, these are not displayed in Figure 4. While associations for these PRSs were generally stronger for ABCD fMRI phenotypes relative to non-fMRI IDPs in ABCD, even the strongest associations were fairly weak (.001 < p < .0002). The only exception was observed for the secondary cluster representing resting-state fMRI IDPs capturing visual cortex signal and interhemispheric connectivity (RVI). Despite being constructed from only 13 IDPs from the UK Biobank, this cluster demonstrated strong (min p=1.21×10^-11^) and widespread correlations with hundreds of ABCD resting state fMRI IDPs.

### Identification and consolidation of independent loci

A total of 413 independent genome-wide significant loci (**Supplementary Table 3a**) were identified across 23 DIMPLE-GWAS clusters; no significant SNPs were detected for two of the secondary resting state fMRI subclusters. Of these loci, 189 were unique to a single cluster (**Supplementary Table** 3b), with the remainder shared across multiple clusters, highlighting the genetic pleiotropy underlying brain imaging-derived phenotypes (Supplementary Table 3c). For instance, the locus at chromosome 1 (153,755,451–156,615,114bp), was significant in clusters related to white matter — including White Matter Connectivity (WMC), General White Matter Tract (WMT), and Cortical Grey-White Intensities (CGW) — indicating strong cross-cluster relevance to white matter structure; several other loci also seemed to map to multiple white matter clusters. Additionally, a well-studied region at chromosome 17q21.31 (44,330,752–44,906,949bp) was shared among multiple gray and white matter clusters spanning diverse brain systems.

### Identification of novel loci

To identify potential novel loci, we compared our cluster-specific GWAS results with lead SNPs from the BIG-40 database using their "Table of local-peak associations" (−log₁₀ P > 7.5). Due to the absence of complete locus boundaries in BIG-40, we employed a conservative approach: loci in our study were classified as novel if they did not encompass any overlapping lead SNP from the BIG-40 table for the corresponding cluster’s constituent IDPs. This analysis identified 104 novel loci across 23 DIMPLE-GWAS clusters (Supplementary Table 3d). As with the heritability analysis above, the identification of multiple novel loci across the clusters suggests that DIMPLE-GWAS approach may capture additional genetic signal not detected in previous single-phenotype analyses.

### Developmental stage analysis

Using MAGMA gene-property analysis of summary statistics derived from the 25 DIMPLE-GWAS clusters, we performed temporal gene expression enrichment analysis against a reference set of 524 samples from the developing human brain, categorized into 11 developmental stages^15,16^. Eight associations survived FDR correction (q < 0.05) across 275 tests (25 clusters × 11 developmental stages). The developmental stage analysis revealed strong prenatal enrichment patterns for most of the structural neuroimaging clusters, with the ventral stream volume cluster showing the most robust prenatal enrichment (**Figure 5**; Supplementary Table 4). By contrast, resting state fMRI network clusters showed distinct temporal profiles, with enrichment in varying postnatal stages. White matter tract clusters demonstrated significant late prenatal enrichment, including WMT (p = 4.51×10⁻L, q = 0.038, β = 0.062) and WMC (p = 1.43×10⁻³, q = 0.049, β = 0.056). Notably, the cortical grey-white intensity cluster (CGW) showed significant enrichment during adolescence (p = 9.93×10⁻L, q = 0.039, β = 0.078), consistent with relatively delayed maturation of cortical microstructural properties.

**Figure 5.**
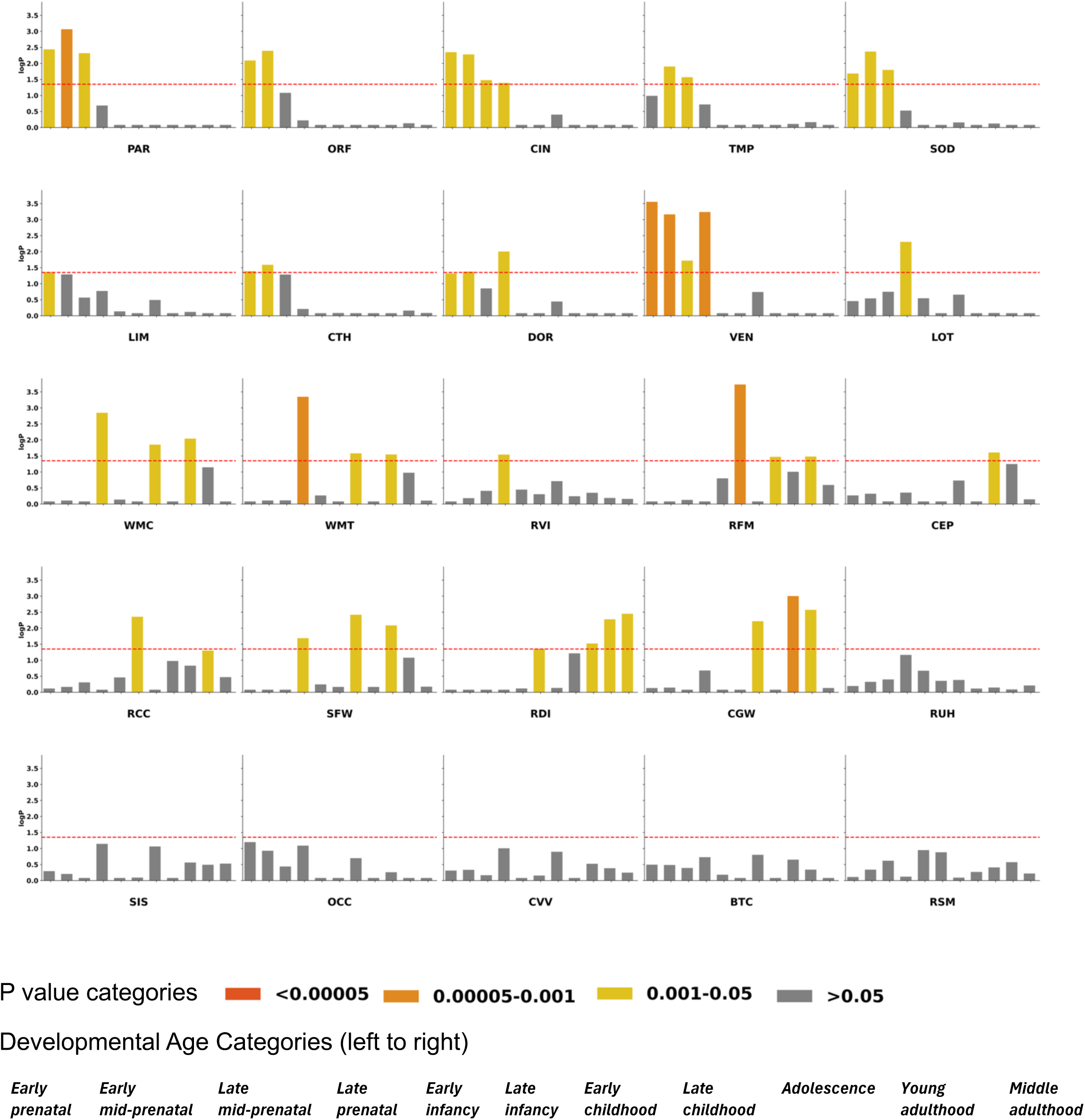
Temporal gene expression enrichment of DIMPLE-GWAS clusters across human brain developmental stages. Heatmap displaying MAGMA-based gene expression enrichment results for each of the 25 DIMPLE-GWAS clusters across 11 developmental age windows spanning early prenatal development through middle adulthood, derived from the BrainSpan atlas. Colour intensity encodes the degree of enrichment (β coefficient), and statistical significance thresholds are indicated as follows: PL<L0.00005; PL<L0.00005-0.001; PL<L0.001-05, P>0.5. Developmental age categories (left to right) are: Early Prenatal, Early Mid-Prenatal, Late Mid-Prenatal, Late Prenatal, Early Infancy, Late Infancy, Early Childhood, Late Childhood, Adolescence, Young Adulthood, and Middle Adulthood. Clusters are grouped by imaging modality. Distinct temporal enrichment signatures across cluster types highlight modality-specific neurodevelopmental trajectories underlying the genetic architecture of brain imaging phenotypes.

### Cell type–specific expression analysis

To resolve the specific cellular contexts of the genetic signals underlying our neuroimaging clusters, we performed cell type–specific expression analysis on the cluster summary statistics, using a reference panel derived from single-nucleus RNA sequencing data from 461 transcriptomically defined cell types across the adult human brain^17,18^. The CGW cluster showed the most robust associations, with 12 Bonferroni-corrected (25 DIMPLE-GWAS clusters * 461 cell types; p<4.34*10^-6^) hits, overwhelmingly implicating the oligodendrocyte lineage (Supplementary Table 5). At a less conservative threshold (25 DIMPLE-GWAS clusters * 31 “superclusters” of cell types; p<6.45*10^-5^), astrocytes also showed enrichment for this cluster. At this lower threshold, enrichment for oligodendrocytes and astrocytes was also observed for several other DIMPLE-GWAS clusters related to white matter phenotypes (CEP, SOD, SFW, and WMT).

### Multiomics association analysis

To identify molecular effects of specific genes underpinning our DIMPLE-GWAS clusters, we performed eQTL, mQTL, sQTL, and pQTL analyses on our cluster summary statistics, resulting in 1,583 genes significantly associated with one or more clusters in at least one molecular modality following multiple testing correction (full results in **Supplementary Table 6**). Gene set enrichment analyses using gProfiler yielded results that differentiated biology underlying the DIMPLE-GWAS clusters, while identifying novel links between several others (**Supplementary Table 7a**). As summarized in **Figure 6**, monoamine transport genes were exclusively associated with a resting state functional MRI cluster containing IDPs representing unimodal-heteromodal connectivity (RUH). Actin and microtubule gene set enrichment was exclusive to the cortical thickness cluster (CTH), while postsynaptic gene set enrichment linked CTH with cingulate volume (CIN). Glial and astrocyte differentiation gene sets were robustly associated with the white matter connectivity (WMC) cluster, but also surprisingly demonstrated a nominally significant association with limbic volume (LIM). Multiple gene sets related to early embryonic development were strongly and exclusively associated with the occipital cortex volume/SA cluster (OCC). By contrast, gene sets specific to neurodevelopmental processes demonstrated widespread associations across white matter, cortical thickness, cortical volume, and subcortical volumetric clusters.

**Figure 6.**
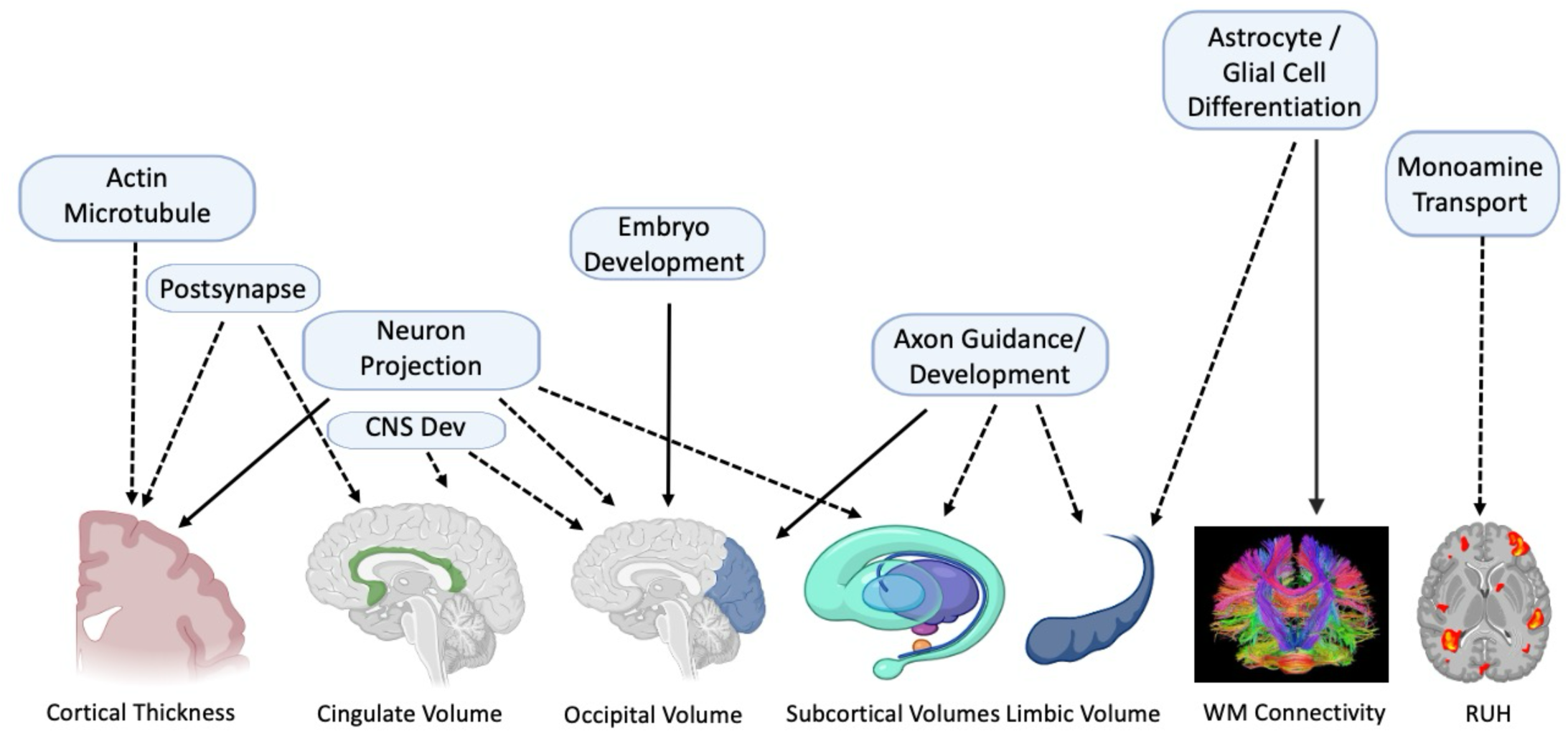
Biological pathway enrichment across neuroimaging genetic architectures. Summary of gene set enrichment analyses characterising the key biological themes underlying each DIMPLE-GWAS cluster that includes synaptic transmission, myelination, neuronal development, and immune-related processes, reflecting the diverse molecular mechanisms underlying genetically distinct brain imaging phenotypes. Pathways were identified through multi-omics integration, including genes implicated via expression (eQTL), methylation (meQTL), splicing (sQTL), and protein (pQTL) quantitative trait loci analyses. Solid arrows represent pathways with FDR-corrected significance; dotted lines represent nominally significant pathways.

As shown in **Supplementary Figure 1**, a total of 166 genes were associated with 3 or more clusters; these were labelled as hub genes. The strongest gene set enrichment for these hub genes was observed for central nervous system development (GO:0007417; adjusted p=2.36×10^-11^). As listed in **Supplementary Table 7b**, approximately one quarter of all significant gene sets were related to anatomical development of neuronal and non-neuronal systems. Axon guidance and glutamatergic synapse gene sets were significantly enriched amongst the hub genes, but other neurotransmitter gene sets were not. Notably, results were not significantly altered when the large group of neighboring genes from the chromosome 17q21 region were removed from the analysis (**Supplementary Table 7c**).

### Association of clusters to observable phenotypes

We estimated pairwise genetic correlations between the 25 DIMPLE-GWAS clusters and 31 phenotypes spanning psychiatric, neurological, personality, behavioral, and cognitive traits using LDSC. After FDR correction for multiple testing across 775 comparisons (25 clusters × 31 traits), 16 correlations achieved significance (P_FDR_ < 0.05) (**Table 1; Supplementary Table 8**). The large cluster comprising all the resting state IDPs (RFM) showed the most robust associations with extraversion (r_g_ = 0.270; P_FDR_ = 3.72×10⁻L), as did the resting state motor network (RSM) sub-cluster (r_g_ = 0.268 P_FDR_ = 0.0155) and the resting-state cortico-cerebellar cluster (RCC) (r_g_ = -0.250; P_FDR_ = 0.0364). RSM was also significantly correlated with risk tolerance (r_g_ = 0.201; P_FDR_ = 0.0471). WMT demonstrated a significant correlation with stroke susceptibility (r_g_ = 0.212; P_FDR_ = 4.61×10⁻L). Corpus callosum/fornix/cingulate connectivity was positively correlated with reaction time (r_g_ = 0.138; P_FDR_ = 9.78×10⁻L), while subcortical volume and subcortical intensity clusters showed negative correlations (r_g_ = -0.137; P_FDR_=0.007 and r_g_ = -0.109; P_FDR_=0.03 respectively). Educational attainment was positively correlated with orbital frontal/insula (r_g_ = 0.096; P_FDR_ = 0.0073) and ventral stream volume (r_g_ = 0.102; P_FDR_ = 0.0115). Resting-state connectivity was negatively correlated with schizophrenia (r_g_ = -0.117; P_FDR_ = 0.0454), while temporal gyrus volume showed negative correlation with major depressive disorder (r_g_ = -0.091, P_FDR_ = 0.0464). Finally, occipital volume was positively correlated with epilepsy (r_g_ = 0.181; P_FDR_ = 0.0471), while cortical grey-white intensities showed negative correlation with obesity (r_g_ = -0.092; P_FDR_ = 0.0454).

**Table 1:**
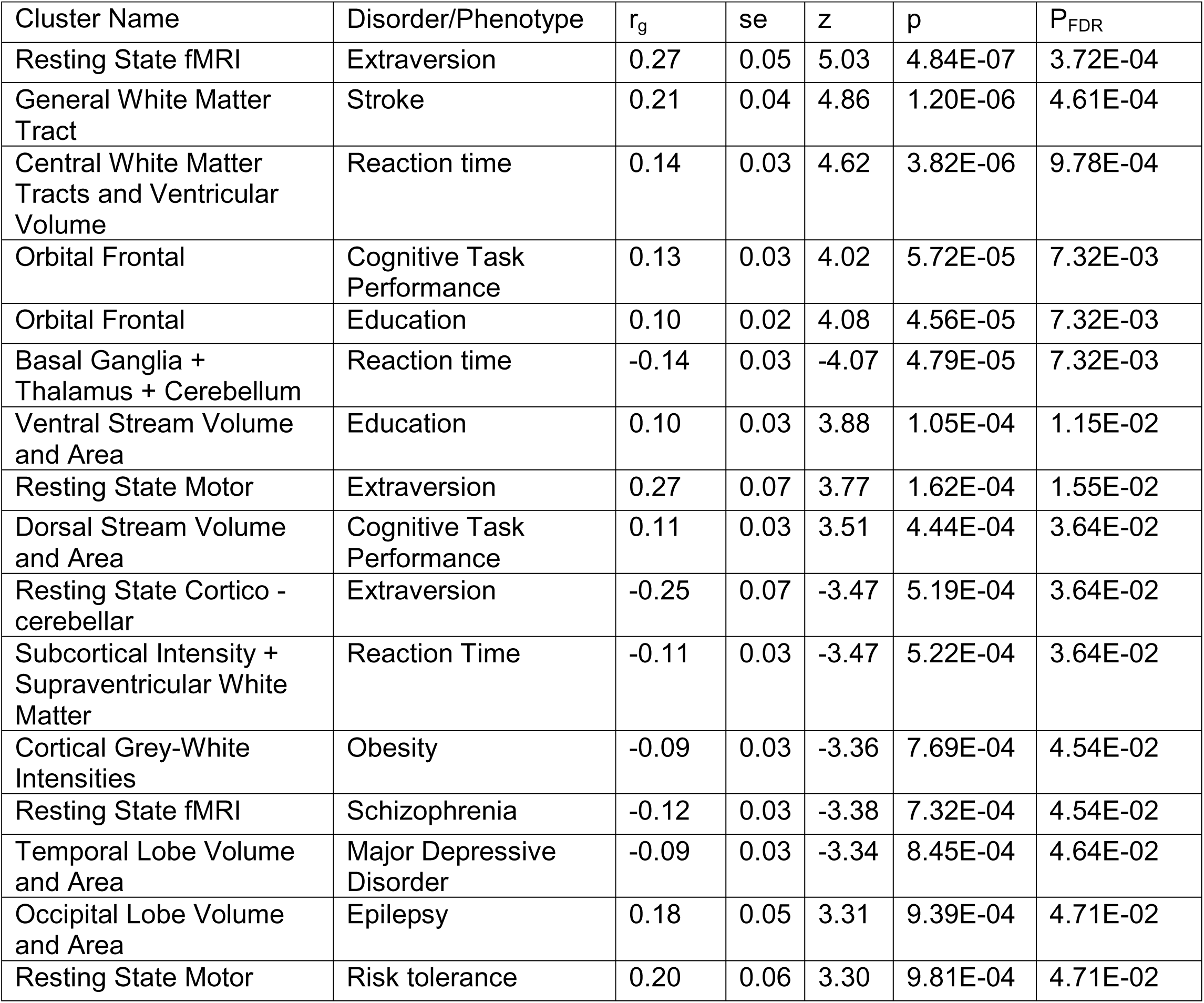
Significant genetic correlations of DIMPLE-GWAS clusters with complex diseases/phenotypes.

| Cluster Name | Disorder/Phenotype | $r_g$ | se | z | p | $P_{FDR}$ |
| --- | --- | --- | --- | --- | --- | --- |
| Resting State fMRI | Extraversion | 0.27 | 0.05 | 5.03 | 4.84E-07 | 3.72E-04 |
| General White Matter Tract | Stroke | 0.21 | 0.04 | 4.86 | 1.20E-06 | 4.61E-04 |
| Central White Matter Tracts and Ventricular Volume | Reaction time | 0.14 | 0.03 | 4.62 | 3.82E-06 | 9.78E-04 |
| Orbital Frontal | Cognitive Task Performance | 0.13 | 0.03 | 4.02 | 5.72E-05 | 7.32E-03 |
| Orbital Frontal | Education | 0.10 | 0.02 | 4.08 | 4.56E-05 | 7.32E-03 |
| Basal Ganglia + Thalamus + Cerebellum | Reaction time | -0.14 | 0.03 | -4.07 | 4.79E-05 | 7.32E-03 |
| Ventral Stream Volume and Area | Education | 0.10 | 0.03 | 3.88 | 1.05E-04 | 1.15E-02 |
| Resting State Motor | Extraversion | 0.27 | 0.07 | 3.77 | 1.62E-04 | 1.55E-02 |
| Dorsal Stream Volume and Area | Cognitive Task Performance | 0.11 | 0.03 | 3.51 | 4.44E-04 | 3.64E-02 |
| Resting State Cortico - cerebellar | Extraversion | -0.25 | 0.07 | -3.47 | 5.19E-04 | 3.64E-02 |
| Subcortical Intensity + Supraventricular White Matter | Reaction Time | -0.11 | 0.03 | -3.47 | 5.22E-04 | 3.64E-02 |
| Cortical Grey-White Intensities | Obesity | -0.09 | 0.03 | -3.36 | 7.69E-04 | 4.54E-02 |
| Resting State fMRI | Schizophrenia | -0.12 | 0.03 | -3.38 | 7.32E-04 | 4.54E-02 |
| Temporal Lobe Volume and Area | Major Depressive Disorder | -0.09 | 0.03 | -3.34 | 8.45E-04 | 4.64E-02 |
| Occipital Lobe Volume and Area | Epilepsy | 0.18 | 0.05 | 3.31 | 9.39E-04 | 4.71E-02 |
| Resting State Motor | Risk tolerance | 0.20 | 0.06 | 3.30 | 9.81E-04 | 4.71E-02 |

As a complementary approach to examining the relationship of the imaging clusters to behavioral phenotypes, we performed two-sample Mendelian randomization analysis using significant SNPs for the DIMPLE-GWAS clusters as exposures and the 31 neurological, psychiatric, and cognitive traits as outcomes. After filtering to exclude associations marked by significant horizontal pleiotropy (Hpleiotropy pval < 0.05) or significant heterogeneity (Heterogeneity pval < 0.05 or extreme outlier values on forest plots), 8 robust (Inverse Variance Weighted FDR<0.05, with weighted median p<0.05) causal associations were observed (**Table 2; Supplementary Table 9).** Notably, the cluster containing subcortical intensity measures (SIS) was causally associated with three psychiatric phenotypes with differing direction of effects: top SNPs for this DIMPLE-GWAS cluster were associated with increasing risk schizophrenia (P_FDR_ = 0.0061) but demonstrated protective effects against anorexia (P_FDR_ = 0.0178) and anxiety disorders (P_FDR_ = 0.0412). Limbic volumes were causally associated with risk for Parkinson’s disease (P_FDR_ = 0.0133). As previously reported^19^, resting-state fMRI showed a causal relationship with insomnia (P_FDR_ = 0.0437); smaller dorsal stream volumes were also associated with increased risk for insomnia (P_FDR_ = 0.0052). White matter subclusters demonstrated two significant associations: long (medial lemniscus/corticospinal) tracts with obesity risk (P_FDR_ = 0.0078), and cerebellar peduncles with cognitive task performance (P_FDR_ = 0.0005).

**Table 2:**
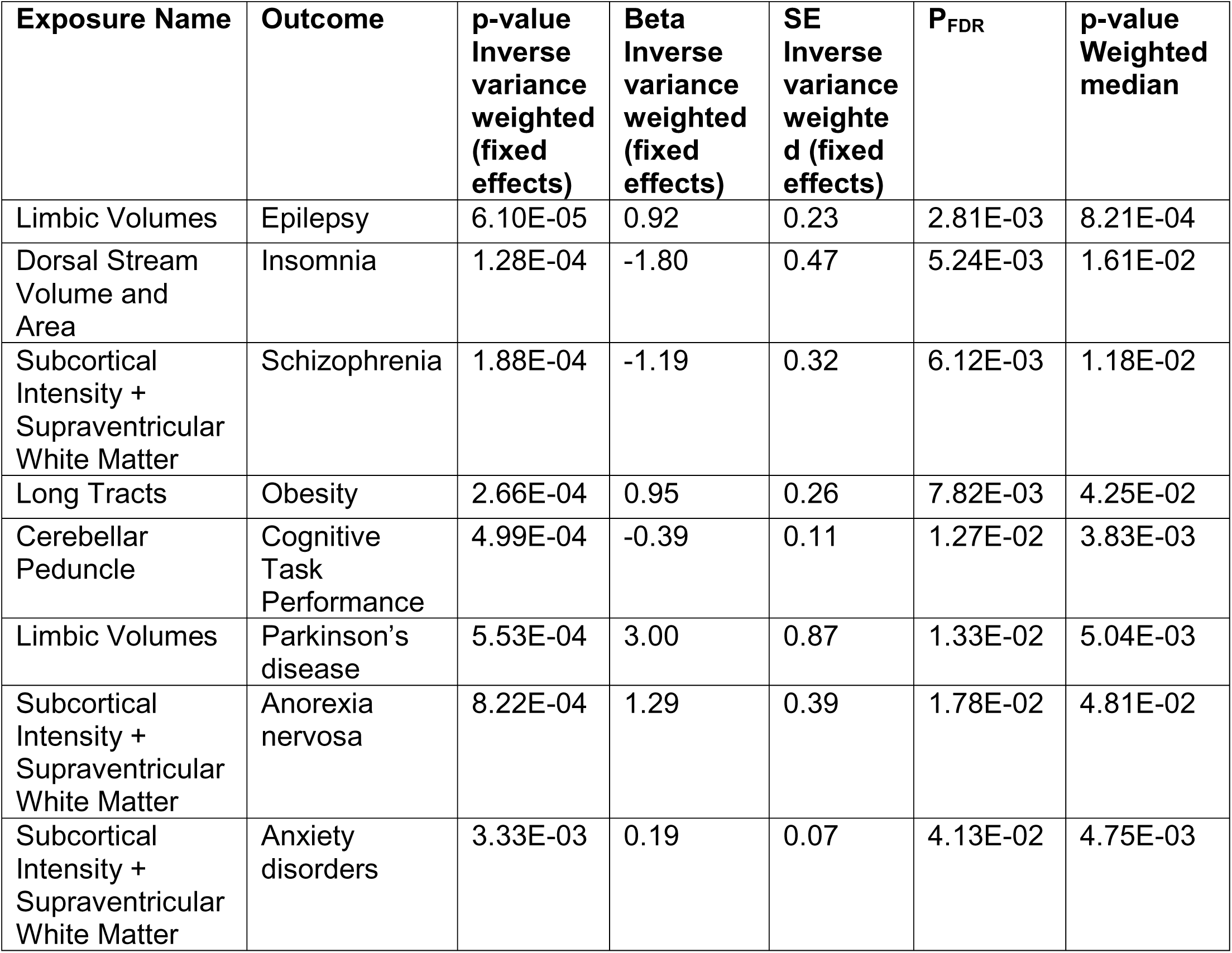
Significant Two-sample Mendelian randomization (MR) output of DIMPLE-GWAS clusters and behavioral phenotypes.

| **Exposure Name** | **Outcome** | **p-value**<br>**Inverse**<br>**variance**<br>**weighted**<br>**(fixed**<br>**effects)** | **Beta**<br>**Inverse**<br>**variance**<br>**weighted**<br>**(fixed**<br>**effects)** | **SE**<br>**Inverse**<br>**variance**<br>**weighted**<br>**(fixed**<br>**effects)** | **P<sub>FDR</sub>** | **p-value**<br>**Weighted**<br>**median** |
| --- | --- | --- | --- | --- | --- | --- |
| Limbic Volumes | Epilepsy | 6.10E-05 | 0.92 | 0.23 | 2.81E-03 | 8.21E-04 |
| Dorsal Stream<br>Volume and<br>Area | Insomnia | 1.28E-04 | -1.80 | 0.47 | 5.24E-03 | 1.61E-02 |
| Subcortical<br>Intensity +<br>Supraventricular<br>White Matter | Schizophrenia | 1.88E-04 | -1.19 | 0.32 | 6.12E-03 | 1.18E-02 |
| Long Tracts | Obesity | 2.66E-04 | 0.95 | 0.26 | 7.82E-03 | 4.25E-02 |
| Cerebellar<br>Peduncle | Cognitive<br>Task<br>Performance | 4.99E-04 | -0.39 | 0.11 | 1.27E-02 | 3.83E-03 |
| Limbic Volumes | Parkinson's<br>disease | 5.53E-04 | 3.00 | 0.87 | 1.33E-02 | 5.04E-03 |
| Subcortical<br>Intensity +<br>Supraventricular<br>White Matter | Anorexia<br>nervosa | 8.22E-04 | 1.29 | 0.39 | 1.78E-02 | 4.81E-02 |
| Subcortical<br>Intensity +<br>Supraventricular<br>White Matter | Anxiety<br>disorders | 3.33E-03 | 0.19 | 0.07 | 4.13E-02 | 4.75E-03 |

## DISCUSSION

With the advent of biobank-scale genomic data, evidence of widespread pleiotropy^20^ has fueled efforts to develop dimensionality reduction approaches to GWAS results. Prior approaches have been largely focused on identify underlying latent phenotypic dimensions, based either on factor analytic^21^(FA) or structural equation modelling^22^ (SEM) methodologies, but these have yielded only limited insights when applied to neuroimaging phenotypes^23–29^, either due to limited input data (as a result of computational limitations in dealing with large numbers of IDPs), or challenges in resolving interpretable latent structures. Here, we introduce a complementary approach, DIMPLE-GWAS, that assigned thousands of multimodal IDPs to latent clusters in a reduced dimensional space. The principles of DIMPLE-GWAS were developed using ground-truth data, and the resulting latent brain endophenotypes were validated in the independent ABCD dataset. While DIMPLE-GWAS was developed in the context of brain IDPs, which tend to demonstrate very high intercorrelations^3,30^ suitable for identification of clusters, the source code for DIMPLE-GWAS will be made freely available so that future work can examine its effectiveness with other pleiotropic phenotypes.

An important goal of DIMPLE-GWAS, as with other dimensionality reduction approaches, is to reduce noise and enhance power for genotype-phenotype associations. Our results demonstrate that the neuroimaging clusters emerging from DIMPLE-GWAS were marked by substantially greater heritability compared to individual IDPs, yielding greater power for locus discovery, including 104 genomewide-significant loci not reported in prior neuroimaging genomics studies. Importantly, these latent phenotypes do not correspond to commonly used brain atlases, distinguishing our results from other recent studies that attempt to combine genetic and neuroanatomic information^6,31^.

Using only genetic covariance information, our results identified several noteworthy distinctions: First, as has been previously demonstrated^24,30,32^, cortical thickness and area measures have distinct underpinnings in genetic variation, with cortical thickness forming a relatively coherent single cluster. By contrast, cortical area measures clustered into discrete regional clusters, primarily divided by gross lobar neuroanatomy rather than more refined gyral^33^ or cytoarchitectonic^34^ distinctions favored by traditional atlases, and only partially reflecting previously identified transcriptional axes of neurodevelopment^35^. While the DIMPLE-GWAS clusters are broadly comparable to results previously reported using FA^26^ or genomic SEM^32^ approaches, the present study utilized a much larger and more regionally refined set of input IDPs, permitting greater clarity and resolution of anatomic boundaries. For example, for the first time we are able to observe clear separation of ventral and dorsal cortical streams^36^, indicating that these classically defined “perception” and “action” pathways^37^ have differential genetic underpinnings. Moreover, we demonstrate that the global measures of cortical area and volume load onto the dorsal cluster, consistent with prior observations derived from strictly neuroanatomic data indicating that total cortical volume is a function of areal expansion (in both evolutionary^38^ and neurodevelopmental^39^ terms) specific to dorsal fronto-parietal cortex, and that global cortical volumes are genetically correlated with dorsal regional volumes^40^. It is important to note that, for each cortical region, area and volume measures co-clustered in DIMPLE-GWAS, despite volume being a function of both area and thickness.

While prior studies of pleiotropy and dimensionality reduction have focused on cortical structural measures such as thickness and area, none have also included signal intensity measures, subcortical volumes, diffusion MRI phenotypes, and functional MRI IDPs as well. DIMPLE-GWAS indicated that cortical signal intensity measures had different genetic associations compared to subcortical signal intensity. Cerebellar volumes clustered with other subcortical structures such as basal ganglia and thalamus, consistent with their neurodevelopmental coordination^41,42^ and extensive monosynaptic and disynaptic links^43–45^, but these were well-differentiated from the limbic (hippocampal/amygdala) cluster. Interestingly, ventricular volumes clustered with diffusion MRI measures of central white matter tracts, and other dMRI IDPs were differentiated by both tract type (e.g., long tracts vs supraventricular) and measurement parameter (e.g., free water vs orientation dispersion). These genetically-defined white matter features differ substantially from developmentally^46^ or phenotypically^47^ defined axes. Functional MRI measures (mostly from resting-state, as task-based fMRI generally did not meet minimal heritability criteria for inclusion in DIMPLE-GWAS) clustered together, with less clear differentiation into subclusters. However, it is noteworthy that the subclusters did not generally recapitulate canonical functional networks^48^, with the exception of the default mode network; it is possible that prior studies demonstrating relatively weak GWAS signals for canonical networks^49,50^ indicate that genetic variation is orthogonal to phenotypic variation across these measures.

Downstream analyses of the summary statistics from the DIMPLE-GWAS clusters revealed several important biological insights. In general, cortical volume/area measures (especially the ventral stream cluster) were enriched for genes expressed prenatally, while resting state clusters were enriched for late-expressed genes and white matter clusters were intermediate between these. Functional gene set enrichment, as summarized in Figure 6, demonstrated surprising links across clusters; for example, global cortical thickness and cingulate volume shared enrichment for postsynaptic gene sets. Additionally, we identified a set of “hub genes,” enriched for neurodevelopmental processes, that were associated across multiple (≥3) clusters. In the supplementary tables, we provide extensive lists of genes with transcriptomic and/or proteomic associations to each cluster, extending recent studies with less comprehensive datasets^51,52^. Examination of associations with other neuropsychiatric, cognitive, and behavioral phenotypes revealed known links, such as white matter with stroke^53^, and novel connections between subcortical measures and reaction time. The protective effects identified, such as dorsal stream volume against insomnia and internal capsule integrity against depression, offer initial targets for future neuroprotective strategies.

In summary, the present study is the first, to our knowledge, to apply UMAP and HDBSCAN to cluster large numbers of GWAS summary statistics to identify latent endophenotypes. It is noteworthy that a recent study applied similar techniques to cluster individual-level genotype data for purposes of identifying ancestry subgroups^53^, suggesting a growing recognition that these methods are well-suited to large-scale genetic data. Despite these advances, several limitations require acknowledgment. The analyses were restricted to European-ancestry populations, limiting generalizability to other groups and potentially missing ancestrally specific loci or genetic architectures. The current GWAS and neuroimaging data remain cross-sectional, complicating the investigation of developmental trajectories or dynamic brain changes. While clustering improves power and interpretability, decisions regarding parameterization and granularity may impact cluster resolution and biological specificity.

## METHODS

### IDP data curation

We curated genome-wide association study (GWAS) summary statistics for 3935 imaging-derived phenotypes (IDPs) from the Oxford Brain Imaging Genetics (BIG-40)^4^ database^54^. These GWAS data were derived from brain MRI-based phenotypes across two UK Biobank (UKBB) releases, comprising approximately 33000 participants of European ancestry. These curated IDPs spanned multiple neuroimaging modalities, including T1-weighted structural MRI, diffusion MRI, susceptibility-weighted imaging (SWI), fluid-attenuated inversion recovery (FLAIR), task-based fMRI, and quality control (QC) metrics.

To ensure robust genetic associations, IDPs for which the h² p-value exceeded 0.05 (i.e., heritability not significantly different from zero) were excluded, refining the dataset to a final set of 2,315 IDPs for downstream analyses. We then extracted approximately 1.2 million HapMap3 SNPs, applying a pruning threshold of r^2^ > 0.1 to linkage disequilibrium (LD) to reduce redundancy, ultimately resulting in a 2,315 x 86,885 IDP x SNP matrix for dimensionality reduction and clustering procedures.

### Clustering using UMAP

The dimensionality reduction and clustering approach implemented in the DIMPLE-GWAS workflow leverages a combination of uniform manifold approximation and projection (UMAP) for non-linear dimensionality reduction and hierarchical density-based spatial clustering of applications with noise (HDBSCAN) for identifying clusters in high-dimensional space. Initially, input data—a matrix of Z-scores from GWAS summary statistics across HapMap3 SNPs—is loaded and transposed, discarding initial metadata columns to focus on genetic signal variance. UMAP, a manifold learning technique grounded in Riemannian geometry and algebraic topology, embeds the high-dimensional data (thousands of IDPs by tens of thousands of SNPs) into a lower-dimensional space (default n_components=3) while preserving both local and global topological structures through a fuzzy simplicial set representation. Mathematically, UMAP optimizes a cross-entropy loss between high-dimensional fuzzy topological representations (constructed via nearest-neighbor graphs with parameters: n_neighbors=30 for local radius and min_dist=0.0 for embedding compactness) and their low-dimensional counterparts, often incorporating densMAP for density-preserving projections. To address UMAP’s inherent stochasticity—arising from random initializations and approximations in the optimization process—the code executes 1,000 parallel UMAP runs with distinct random seeds (1 to 1,000), stacking the resulting embeddings into a concatenated matrix (e.g., 3,000 dimensions), thereby capturing ensemble variations and enhancing robustness for downstream clustering.

Following embedding, HDBSCAN performs density-based clustering without presupposing cluster count, modeling clusters as regions of high density separated by low-density areas via a hierarchical approach: it constructs a minimum spanning tree from mutual reachability distances (defined as max(core_k_distance(p), core_k_distance(q), d(p,q)) where core_k_distance is the distance to the k-th nearest neighbor, with k=min_samples), then condenses it into a hierarchy pruned by stability criteria using parameters including min_cluster_size (minimum points per cluster) and cluster_selection_epsilon (threshold for merging clusters in the ’eom’ method). A parallelized grid search exhaustively evaluates combinations across min_samples (2-20), min_cluster_size (5-50), and epsilon (0-3), filtering for configurations yielding >1 cluster and computing metrics such as the average silhouette score (ranging from -1 to 1, measuring intra-cluster cohesion versus inter-cluster separation via s(i) = (b(i) - a(i)) / max(a(i), b(i)) where a(i) is average distance to same-cluster points and b(i) to nearest-cluster points). Valid configurations are ranked using a weighted scoring formula balancing silhouette score, cluster count, and noise proportion (number of unclustered datapoints). Clustering quality is further assessed with the Calinski-Harabasz index (variance ratio criterion, maximizing between-cluster dispersion over within-cluster dispersion), Davies-Bouldin index (average similarity ratio of clusters, minimizing intra-cluster distances relative to inter-cluster centroids), and Dunn index (ratio of minimum inter-cluster distance to maximum intra-cluster diameter). Embedding fidelity is validated via trustworthiness (1 - (2 / (n * k * (2n - 3k - 1))) * sum(V), penalizing high-dimensional neighbors poorly preserved in low dimensions) and continuity (symmetric measure for low-dimensional neighbors absent in high dimensions), both using k-nearest neighbors (default k=30). The workflow culminates in interactive 3D visualizations using Plotly for cluster exploration (color-coded by labels or modalities) and static silhouette plots with Matplotlib, with outputs saved as CSV for parameter grids and HTML/PNG for figures, enabling reproducible pleiotropic pattern discovery.

### Clustering optimisation and parameter selection

To avoid subjective selection among numerous plausible HDBSCAN solutions, we performed automated optimisation over a hyperparameter grid spanning cluster_selection_epsilon (ε), min_samples, and min_cluster_size. Each candidate solution was summarised using three diagnostics: (i) average silhouette score (cluster separation), (ii) the number of clusters detected (solution granularity; excluding noise), and (iii) the number of points labelled as noise (unassigned observations). Each metric was normalised to a common 0–1 scale, weighted according to its dynamic range across the grid, and combined into a single objective function that rewards higher silhouette and higher cluster count while penalizing noise; the top-ranked solution was then selected for downstream interpretation. For interpretability, ranked objective values were additionally visualised as a fit score curve, with the best model anchored near 1 and poorer solutions approaching 0.

### Genomic PCA meta-analysis

We performed genetic principal component analysis (PCA) on each cluster of imaging-derived phenotypes (IDPs) using a correlation-based framework^11^. Summary statistics for all the IDPs were reformatted using munge_sumstats.py from LDSC for pairwise genetic correlations analysis between IDPs within a cluster using linkage disequilibrium score regression (LDSC). A genetic correlation matrix was constructed for each cluster, adjusting for sample overlap via LDSC intercepts^17,55^. LDSC analysis was restricted to HapMap3 SNPs.

The genetic correlation matrix underwent eigen decomposition to extract eigenvectors and eigenvalues. The first principal component (PC), explaining the largest proportion of shared genetic variance, was retained. Eigenvectors were standardized by scaling with the square root of their corresponding eigenvalues, yielding interpretable weights that reflect the relative contribution of each IDP to the genetic PC. We adapted the multivariate Genome-Wide Association Meta-Analysis (GWAMA) framework^56^ to integrate directional loadings and generate genetic PC summary statistics for each cluster. This approach allowed for the aggregation of SNP effects, optimizing the representation of shared genetic architecture among the IDPs. By focusing on genetic correlation patterns rather than scale-dependent variance, this method facilitated a robust meta-analysis of neuroimaging traits with heterogeneous measurement units within each cluster.

### Heritability analysis

To evaluate the genetic utility of HDBSCAN-UMAP derived clusters, we compared cluster-level heritability estimates against individual IDP heritabilities within each cluster. For each of the 25 clusters, SNP-based heritability (h²) was estimated using linkage disequilibrium score regression (LDSC) ^17,55^. Individual IDP heritability estimates were obtained from previous publications^4^. LDSC analyses used European ancestry LD scores from the 1000 Genomes Project reference panel and only selected HapMap3 SNPs with INFO ≥ 0.7. Enhancement was defined as cluster h² exceeding the mean individual IDP h² within that cluster, indicating improved genetic signal capture through dimensionality reduction of functionally related brain phenotypes.

### PRS validation with ABCD data

To validate the reproducibility of IDP-based clusters derived from DIMPLE-GWAS, polygenic risk scores (PRS) were calculated (using PRS-CS^57^ software with default parameters) in unrelated European-ancestry individuals from the Adolescent Brain Cognitive Development (ABCD) Study^58^ baseline cohort. GWAS summary statistics from the 25 cluster-based DIMPLE-GWAS analyses served as the discovery datasets for computing PRS weights. Details of the processing of ABCD genotyping data^59^ and neuroimaging data^14^ have been reported elsewhere. We hypothesized that if the DIMPLE-GWAS clusters showed associations with PRS for similar IDPs in the ABCD cohort, which would serve as evidence that our clustering approach accurately identified groups of IDPs with shared genetic architecture rather than reflecting random patterns. Linear models were fitted for each of the 25 cluster-based PRS (IDP ∼ PRS + sex + age + site + 10 principal components).

### Neurological/psychiatric GWAS data curation

We systematically curated GWAS summary statistics for various neurological, psychiatric, and cognitive/behavioral traits to evaluate their genetic correlations with neuroimaging-derived clusters using LD score regression (LDSC) and to serve as outcomes in our subsequent two-sample Mendelian randomization analyses.

We curated 11 psychiatric phenotypes, which includes schizophrenia^60^ (SCZ), bipolar disorder^61^ (BIP), major depressive disorder^62,63^ (MDD), attention-deficit/hyperactivity disorder^64^ (ADHD), anxiety disorders^65^ (ANX), autism spectrum disorder^66^ (ASD), post-traumatic stress disorder^67^ (PTSD), and anorexia nervosa^68^ (AN), obsessive-compulsive disorder^69^ (OCD), broad antisocial behavior^70^, and panic disorder^71^ (Supplementary Table 1). We curated 8 Neurological phenotypes that includes including Alzheimer’s disorder^72^ (AD), amyotrophic lateral sclerosis (ALS)^73^, epilepsy^74^, headache^20^, migraine^75^, Parkinson’s disease^76^ (PD), stroke^77^, insomnia^78^ (Supplementary Table 1). Additionally, GWAS datasets for personality, behavioral, and lifestyle traits were included: agreeableness, conscientiousness, extraversion, neuroticism, openness^79^, alcohol consumption per week, cigarettes smoked per day^80^, obesity^81^, and risk tolerance^82^. We also included three cognitive phenotypes that include reaction time^83^, cognitive task performance^84^, and educational attainment^85^ (Supplementary Table 1).

All GWAS datasets were sourced from publicly available consortia or peer-reviewed studies (see Supplementary Table 1). To minimize population stratification, we restricted all GWAS datasets to populations of European ancestry. Additionally, where possible, cohorts excluding UK Biobank (UKBB) participants were prioritized to mitigate potential confounding from sample overlap with the neuroimaging IDP cohort.

### Neurological/psychiatric GWAS data QC

We first converted all neurological, psychiatric, and cognitive GWAS summary statistics obtained from public repositories into VCF format using the gwas-to-vcf tool^86,87^ to harmonize effect allele to alternative allele across datasets. Next, we incorporated alternative allele frequency data from the European ancestry dbSNP cohort using bcftools^88^ (Version: 1.19).

Variants exhibiting substantial discrepancies (>20%) in alternative allele frequency between the GWAS datasets and the dbSNP European ancestry cohort were removed. Additionally, we excluded all insertion-deletion variants, as well as variants with INFO scores below 0.7 or alternative allele frequencies lower than 0.005. We excluded the MHC region (chr6:25Mb-35Mb) from all the clusters due to its complex LD structure.

### Summary statistics imputation

Of the 31 GWAS we have curated for our study, 11 GWAS were relatively deficient in SNP density after QC (less than n=7000000) and therefore we used the summary-gwas-imputation pipeline^89^ with default parameters (1 Mb non-overlapping windows, ridge regression penalty λ = 0.1, and used only variants with MAF >0.001) to harmonize variants to the GTEx v8 reference EUR panel (provided in their previous publication^90^ and impute missing variants for those summary statistics. The script Summary-gwas-imputation pipeline imputes missing summary stats (zscores) using the covariance of the LD reference data set and the existing summary stats, within a specification of a region. The underlying algorithm is the one from previously published methods (DIST and ImpG-Summary/LD)^91,92^, that assume a multivariate Gaussian distribution for the association summary statistics. Zscores created by the summary-gwas-imputation pipeline were converted to beta and SE^93^. Finally, the imputed summary statistics were converted to VCF format using the gwas-to-vcf tool, and all QC steps mentioned above were performed to prepare the data for downstream analyses such as genetic correlation estimation or Mendelian randomization.

### Identification and consolidation of independent loci

Independent genome-wide significant loci (p<5×10^-8^) were identified and clumped using Functional Mapping and Annotation (FUMA; version 1.5.2) for each of the 25 clusters separately. The detailed FUMA pipeline was described previously^94^ Briefly, it employs a two-step clumping process based on reference data from the European 1000 Genomes Project (Phase 3) (8). At first, SNPs were clumped based on a p-value < 5×10^-8^ and an LD threshold of r^2^ < 0.6 to identify “independent significant SNPs.” A second clumping was performed to identify “lead SNPs” (the strongest signal at a given locus) using a tighter LD threshold of r^2^ < 0.1. SNPs within 250 kb were merged into single genomic loci for each cluster^94^.

### Identification of novel loci

We attempted to identify if our study identified any new loci. As direct locus-to-locus comparisons were infeasible due to the absence of full locus boundaries in BIG-40^4^ database^54^, we utilized the "Table of local-peak associations (-Log10(P) > 7.5)" provided by BIG-40, which catalogs lead SNPs surpassing this stringent significance threshold. We have extracted the lead SNPs for each cluster based on the IDPs included in those clusters. For each cluster, we extracted lead SNPs associated with constituent IDPs from this table and checked for overlap with loci identified in our cluster GWAS. Loci in our study lacking any overlapping lead SNP from the BIG-40 table were classified as novel.

### Developmental stage analysis

To investigate temporal patterns of gene expression during brain development, we analyzed developmental age-based gene expression profiles using 524 samples from the developing human brain^95^. Samples were categorized into 11 developmental stages and 29 age groups based on post-conceptional weeks, excluding age groups with fewer than 3 samples (25 pcw and 35 pcw). From 52,376 annotated genes, we retained genes with average RPKM >1 in at least one developmental stage or age group, resulting in 19,601 genes for developmental stages and 21,001 genes for age groups. RPKM values were winsorized at 50 and log2-transformed with pseudocount 1. MAGMA gene-property analysis was performed using the average log-transformed expression per developmental label as covariates, conditioning on the average expression across all labels to identify developmentally specific expression patterns associated with our genetic clusters.

### Single cell type analysis

To identify cell type-specific genetic architecture underlying neuroimaging-derived phenotypes (IDPs), we leveraged 461 transcriptomically defined cell types that were curated using snRNA-seq dataset from 3,369,219 nuclei across 105 postmortem brain dissections^18^. For each cluster, we regressed MAGMA-derived gene z-scores against cell type specificity scores in a multivariate framework that accounted for LD between genes and included covariates for gene size, density, sample size, and minor allele frequency. To identify independent cell type associations, we implemented a conditional analysis protocol: using forward stepwise selection, we iteratively evaluated pairs of cell types through proportional significance (PS) metrics. Cell types were retained in the final set if they either: (1) showed mutual independence (PS ≥ 0.8 in both directions), or (2) demonstrated partial independence (0.5 ≤ PS < 0.8) while maintaining marginal significance (p ≤ 0.05). This conservative approach ensured robust identification of cell types with independent contributions to phenotype associations while accounting for potential conditional relationships where less significant cell types might harbor stronger independent signals.

### Molecular association analyses and multi-omics integration

We integrated multiple layers of molecular evidence to link DIMPLE-derived neuroimaging clusters to putative causal genes. All upstream analyses were performed in R, and detailed implementation steps are provided in the Supplementary Methods.

First, we generated modality-specific gene–cluster association maps. For QTL-based analyses, we used summary-based Mendelian randomisation (SMR) with HEIDI heterogeneity testing applied separately to expression (eQTL), splicing (sQTL) and methylation (mQTL) data. For each QTL layer, we retained gene–cluster pairs that passed both an FDR-corrected SMR significance threshold (< 0.05) and a linkage confound filter (p__HEIDI_ ≥ 0.001), and discarded associations with missing or non-interpretable HEIDI statistics.

Second, we incorporated protein-level evidence from Mendelian randomisation of blood and brain protein QTLs. For blood pQTLs, cis and trans instruments were analysed separately, and gene–cluster associations were carried forward if they met an FDR threshold of 0.05. Brain-relevant expression and protein effects were captured using an integrated brain QTL/MR dataset, again retaining only FDR-significant gene–cluster pairs (FDR ≤ 0.05). These QTL and MR analyses yielded a harmonised set of associations annotated by DIMPLE cluster, gene symbol, molecular modality and statistical stringency.

Third, we integrated a complementary, model-based source of evidence using the FLAMES framework, which aggregates genomic, transcriptomic and epigenetic information into gene–cluster precision scores. We restricted this layer to high-confidence FLAMES links with cumulative precision ≥ 0.75. These FLAMES-derived gene–cluster pairs were then harmonised to the same schema as the QTL/MR results.

All molecular inputs were subsequently merged into a single long-format matrix (Overall-omics), in which each row corresponds to one gene–cluster association annotated with its analysis method (SMR, blood pQTL, brain MR, FLAMES) and molecular modality (e.g. eQTL, sQTL, mQTL, cis/trans pQTL, brain pQTL). Within each stream, we removed entries with missing or non-informative cluster labels, applied the modality-specific thresholds described above, and de-duplicated repeated gene–cluster pairs.

From this integrated matrix we quantified, for each gene, the number of distinct DIMPLE clusters to which it was linked (cross-cluster connectivity) and the number of distinct molecular modalities supporting those links. Genes connecting to at least three clusters were defined as multi-omics “hub” genes, while those associated with a single cluster were classified as “singletons”. Hub genes and their cluster connectivity were visualised in a gene–by–cluster dot-plot, where dot size represented the number of supporting modalities and colour encoded the number of connected clusters. In parallel, we constructed a cluster–cluster network in which edges between DIMPLE clusters were weighted by the number of shared hub genes and the diversity of modalities supporting those genes. Together, these complementary views summarise how multi-omics evidence concentrates on specific imaging clusters, identifies cross-cluster hub genes, and highlights densely interconnected regions of the neuroimaging landscape.

### LDSC

We estimated pairwise genetic correlations between each of the 25 phenotype clusters and a broad spectrum of psychiatric, neurological, personality/behavioral, lifestyle, and cognitive traits (Supplementary Table 1) using bivariate linkage disequilibrium score regression (LDSC)^17,55^. For all analyses, we restricted SNPs to the high-quality HapMap3 panel, excluding variants with INFO score <0.7, ambiguous alleles (A/T or C/G) which present strand orientation uncertainty, and multiallelic sites. Intercepts in the regression models were left unconstrained to correct for potential sample overlap and residual population stratification, thus preventing inflation of correlation estimates.

### Two-sample MR Neurological/Psychiatric MR

We performed two-sample MR analyses using the TwoSampleMR software (version 0.6.15)^96^, where top SNPs for each cluster were used as the exposure and neurological, psychiatric, and cognitive trait GWASs were used as outcome. We removed palindromic variants with MAF > 0.42. After the QC-filtering step, we selected variants that achieve genome-wide significance (P < 5 × 10^−8^) from Cluster GWAS for LD pruning (1000 genome EUR as reference panels) to identify strong, independent genetic instruments. Inverse-variance fixed effect method (IVW-fixed effect) tests with default parameters were the primary test, followed by tests of causal direction using the MR Steiger test, horizontal pleiotropy using the MR-Egger intercept test, heterogeneity using Cochran’s *Q* statistic, and sensitivity analyses using the weighted median statistic and the MR-Egger test.

## Supporting information

Supplementary Figure and Table Description

## Data Availability

All data produced in the present work are contained in the manuscript

## Data Availability

GWAS summary statistics for 3,935 neuroimaging-derived phenotypes (IDPs) from the UK Biobank are publicly available through the Oxford Brain Imaging Genetics (BIG-40) server (https://open.win.ox.ac.uk/ukbiobank/big40/). GWAS summary statistics for all 31 psychiatric, neurological, and cognitive/behavioral traits used for LDSC and Mendelian randomization analyses were obtained from publicly available sources; accession details and URLs for each dataset are provided in Supplementary Table 10. Adolescent Brain Cognitive Development (ABCD) Data used for polygenic risk score validation are available through the NIMH Data Archive (https://nda.nih.gov/abcd) available upon request. All relevant instructions to obtain the data can be found online (https://nda.nih.gov/abcd/request-access). Gene expression data for developmental stage enrichment analyses were obtained from the BrainSpan Atlas of the Developing Human Brain (https://brainspan.org). QTL reference datasets used in SMR/HEIDI analyses (eQTL_eQTLGen, eQTL_BrainMeta, sQTL_BrainMeta, sQTL_GTEx_Whole_Blood, mQTL_BrainMeta, mQTL_McRae) are available through the SMR Portal (https://yanglab.westlake.edu.cn/software/smr/). GTEx v8 tissue-specific expression data are available at https://gtexportal.org. We have used publicly available snRNA-seq dataset curated from 3,369,219 nuclei across 105 postmortem brain dissections^18^. We utilized publicly available serum protein pQTL data from UK Biobank Pharma Proteomics Project^97^ and deCODE Genetics^98^. Brain pQTL data for 1,667 proteins from 1,277 post-mortem brains are available publicly^99^. DIMPLE-GWAS cluster-level GWAS summary statistics generated in this study will be deposited in a public repository upon acceptance. The DIMPLE-GWAS source code is available on GitHub (https://github.com/mlamcogent/DIMPLE-GWAS).

## Acknowledgements

U.B., J.J., M.P., T.L., and M.L. were supported by the National Institute of Mental Health of the National Institutes of Health (NIH) under award no. R01MH117646 (T.L., principal investigator). The funding agency had no direct role in the design and conduct of the study; collection, management, analysis, and interpretation of the data; preparation, review, or approval of the manuscript; and decision to submit the manuscript for publication. The content is solely the responsibility of the authors and does not necessarily represent the official views of the NIH.

## Author Contributions

T.L. and M.L. conceived and designed the study. M.L. designed the data analytic workflow. U.B. and J.J. performed the primary data analyses. Y.Z. and T.G. performed the data analysis with ABCD cohort. T.G. also provided intellectual contributions to the data analytic approach. M.P. critically reviewed reporting of methods and results. U.B., J.J., M.L., and T.L. wrote the manuscript, with critical input from all authors. T.L. supervised the study and acquired funding.

