## Supplementary Figure and Table Description for "A Pleiotropic Map of Brain Imaging Genetics Reveals Biologically Distinct Latent Endophenotypes"

**Supplementary Figure 1 | Multi-omic integration identifies highly connected hub genes and cross-cluster network architecture.**

Hub genes are defined as genes robustly associated with three or more distinct DIMPLE-GWAS imaging clusters across at least one multi-omic modality (eQTL, meQTL, sQTL, or pQTL).

a. Gene–cluster dot-plot matrix showing the distribution of hub genes across DIMPLE-GWAS clusters. Genes (y-axis) are ordered in descending order of the number of clusters with which they associate; DIMPLE-GWAS cluster codes are displayed on the x-axis. Point size encodes the number of supporting multi-omic modalities per gene–cluster pair (n_modalities), and point colour intensity (pale to bright red) indicates the total number of distinct clusters linked to each gene (n_clusters). Hub genes appearing across the greatest number of clusters and modalities are positioned at the top of the y-axis, highlighting the most pleiotropically connected loci in the neuroimaging genetic landscape.

b. Cluster–cluster network demonstrating shared genetic architecture across brain imaging phenotypes. Nodes represent individual DIMPLE-GWAS clusters (labelled with three-letter cluster codes). Edges connect pairs of clusters that share at least one hub gene; midpoint dots are sized proportional to the number of shared hub genes and coloured by the diversity of supporting multi-omic modalities (pale to bright red scale). Network layout was generated using the Fruchterman–Reingold force-directed algorithm, such that clusters with greater shared genetic content are positioned in closer proximity. This network reveals modality-spanning hubs of genetic connectivity and identifies putative biological axes that cut across conventionally distinct imaging domains.

**Supplementary Table 1: DIMPLE-GWAS Clustering Results (Stage 1 & 2)**

- Primary and secondary clustering results from the DIMPLE-GWAS pipeline applied to UK Biobank neuroimaging data.
- 1a: Stage 1 Clustering (2,315 IDPs into 15 primary clusters)
- Contains IDP identifiers, descriptions, heritability estimates (Ph² with SE), UMAP coordinates (3 dimensions), and cluster assignments. Silhouette = 0.612, Fit = 0.846.
- 1b: Stage 2 White Matter Subclustering (459 IDPs into 6 clusters)
- Secondary clustering of white matter IDPs with UMAP coordinates in both primary and secondary embedding spaces.
- 1c: Stage 2 fMRI Subclustering (164 IDPs into 5 clusters)
- Secondary clustering of resting-state fMRI IDPs with coordinates in both embedding spaces.
- 1d: Cluster Nomenclature Reference
- Standardized names and 3-letter codes for all 25 clusters (15 primary + 10 secondary). Essential reference for identifying clusters throughout the paper.

**Supplementary Table 2: PRS Validation in ABCD Study**

- Validation of DIMPLE cluster PRSs in independent ABCD cohort (N=4,264 adolescents): association test between UK Biobank-derived cluster PRSs and corresponding imaging phenotypes to validates biological relevance of cluster definitions in independent population.

**Supplementary Table 3: Genome-Wide Significant Loci (GRCh37/hg19)**

- 3a: All Independent Loci (413 loci, p < 5×10⁻⁸)
- Complete catalog across all clusters with chromosome, position, boundaries, lead SNP, all independent significant SNPs, and p-values as identified using FUMA.
- 3b: Cluster-Unique Loci (189 loci)
- Loci showing genome-wide significance in only one cluster.
- 3c: Shared Loci (224 loci)
- Loci showing genome-wide significance across multiple clusters
- 3d: Novel Loci (104 loci)
- Previously unreported loci revealed through DIMPLE clustering approach.

**Supplementary Table 4: Temporal Gene Expression Enrichment**

- MAGMA analysis testing gene expression enrichment across 11 human brain developmental stages (early prenatal through middle adulthood) using BrainSpan atlas data.

**Supplementary Table 5: Cell Type-Specific Genetic Architecture**

- MAGMA analysis linking genetic architecture to 461 adult human brain cell types. Contains 11,525 results (25 clusters × 461 cell types) with beta values, SE, p-values, and comprehensive cell type annotations (supercluster, cell class, neurotransmitter type). Identifies cell-type-specific genetic mechanisms underlying imaging variation.

**Supplementary Table 6: Multi-Omics Mendelian Randomization (a-f)**

- 6a: eQTL SMR (Dataset: eQTL_BrainMeta, eQTL_Gen)
- Summary-data-based Mendelian Randomization (SMR) of expression quantitative trait loci (eQTL) for DIMPLE-GWAS clusters
- 6b: meQTL SMR (Dataset: mQTL_BrainMeta, mQTL_McRae)
- Summary-data-based Mendelian Randomization (SMR) of methylation quantitative trait loci (meQTL) for DIMPLE-GWAS clusters
- 6c: sQTL SMR (Dataset: sQTL_BrainMeta, sQTL_GTEx_Whole_Blood)
- Summary-data-based Mendelian Randomization (SMR) of splicing quantitative trait loci (sQTL) for DIMPLE-GWAS clusters.
- 6d: Cis-pQTL MR (UKB-PPP, DeCODE)
- Two-sample MR of blood proteins with IVW (primary), MR-Egger, weighted median, Wald ratio methods. Includes directionality testing, heterogeneity, and pleiotropy assessment.
- 6e: Trans-pQTL MR (UKB-PPP, DeCODE)
- Trans-acting protein QTLs from blood using comprehensive MR framework.
- 6f: Brain pQTL MR (4,553 proteins, brain)
- Two-sample MR of brain proteins with IVW (primary), MR-Egger, weighted median, Wald ratio methods. Includes directionality testing, heterogeneity, and pleiotropy assessment.

**Supplementary Table 7: Multi-Omics Pathway Enrichment (a-c)**

- 7a: All Multi-Omics Pathways (533 pathways)
- Gene set enrichment across all genes implicated in clusters through eQTL, meQTL, sQTL, or pQTL. Includes database source, adjusted p-value (FDR), pathway size, overlap statistics.
- 7b: Hub Gene Pathways (248 pathways)
- Pathways enriched among 'hub genes' (genes associated with ≥3 clusters). Identifies highly-connected genetic nodes.
- 7c: Hub Gene Pathways Excluding 17q21 (237 pathways)
- Hub gene pathways after excluding the complex 17q21 locus. Shown alongside 7b for before/after comparison.

**Supplementary Table 8: Genetic Correlations via LDSC (a-b)**

- 8a: Cluster-to-Phenotype Correlations
- LDSC genetic correlations between each cluster and psychiatric/neurological/behavioral phenotypes. Includes rg with SE, Z-score, p-value, heritability estimates, intercept, and FDR-corrected p.
- 8b: Pairwise Cluster Correlations
- Genetic correlations among all 25 clusters revealing shared genetic architecture.

**Supplementary Table 9: Two-Sample Mendelian Randomization**

- Two-sample MR testing causal relationships between DIMPLE clusters (exposure) and complex traits (outcome); tested with multiple MR methods (IVW primary, Egger, weighted median, Wald ratio). Includes directionality testing (Steiger filtering), heterogeneity assessment, horizontal pleiotropy testing, and FDR-corrected p-values.

**Supplementary Table 10: Phenotype Metadata and Source Information**

- Reference information for all 31 complex traits/disorders in LDSC and MR analyses. For each phenotype: full name, standardized abbreviation (as used in Tables 8 & 9), PMID, journal, year, and download link to GWAS summary statistics. Enables reproducibility and source verification.
