## Supplementary Figure 1 for "A Pleiotropic Map of Brain Imaging Genetics Reveals Biologically Distinct Latent Endophenotypes"

Supplementary Figure 1 | Multi-omic integration identifies highly connected “hub genes” (defined as genes robustly associated with ≥3 distinct DIMPLE-GWAS imaging clusters) and cross-cluster network architecture.

A, Gene–cluster dot-plot matrix showing the distribution of hub genes across DIMPLE clusters. Genes (y-axis) are ordered in descending order of the number of clusters with which they associate; DIMPLE-GWAS cluster codes are shown on the x-axis. Point size encodes the number of supporting multi-omic modalities per gene–cluster pair (n_modalities), and point colour intensity (pale to bright red) indicates the number of distinct clusters linked to each gene (n_clusters). B, Cluster–cluster network demonstrating the shared genetic architecture across brain imaging phenotypes. Nodes represent individual DIMPLE-GWAS clusters (blue rectangular labels). Edges connect pairs of clusters that share at least one hub gene; midpoint dots are sized proportional to the number of shared hub genes and coloured by the diversity of supporting multi-omic modalities (pale to bright red scale). Layout was generated using the Fruchterman–Reingold force-directed algorithm.

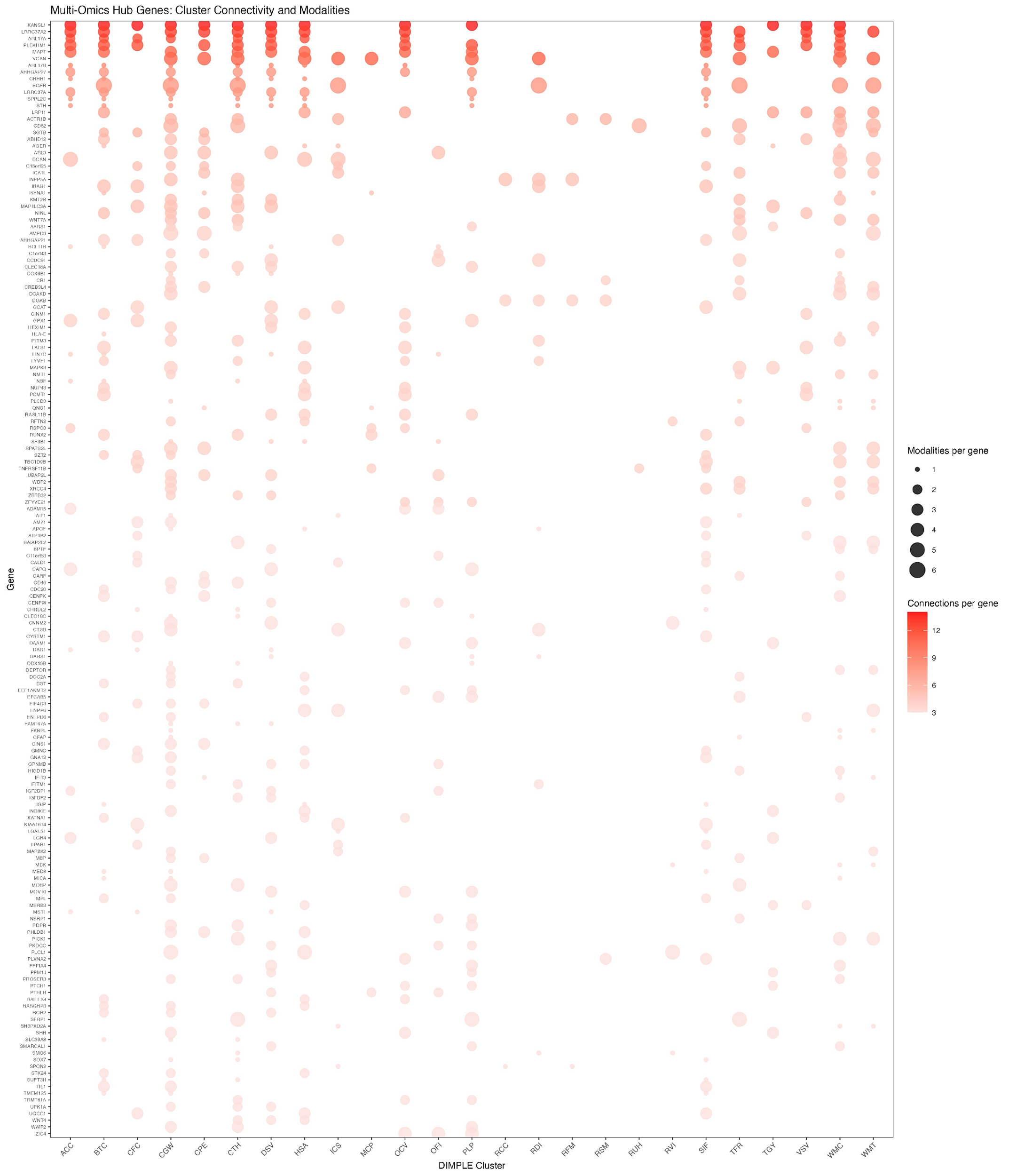

A.

B.

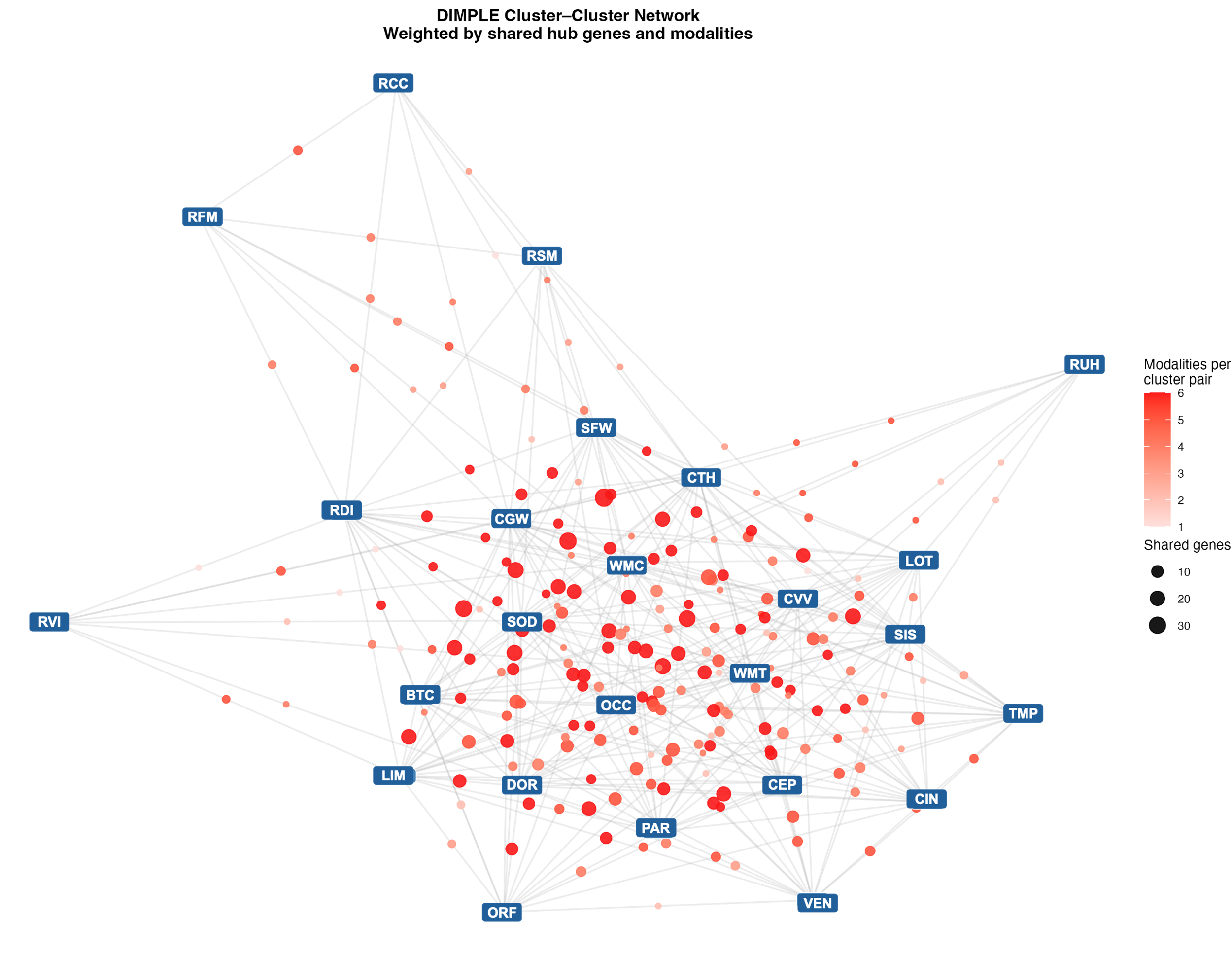
